# A comparative analysis of dengue, chikungunya, and Zika in a pediatric cohort over 18 years

**DOI:** 10.1101/2025.01.06.25320089

**Authors:** Fausto Andres Bustos Carrillo, Sergio Ojeda, Nery Sanchez, Miguel Plazaola, Damaris Collado, Tatiana Miranda, Saira Saborio, Brenda Lopez Mercado, Jairo Carey Monterrey, Sonia Arguello, Lora Campredon, Zijin Chu, Colin J. Carlson, Aubree Gordon, Angel Balmaseda, Guillermina Kuan, Eva Harris

## Abstract

**Background:** Dengue, chikungunya, and Zika are diseases of major human concern. Differential diagnosis is complicated in children and adolescents by their overlapping clinical features (signs, symptoms, and complete blood count results). Few studies have directly compared the three diseases. We aimed to identify distinguishing pediatric characteristics of each disease.

**Methods:** Data were derived from laboratory-confirmed cases (symptomatic infections) aged 2-<18 years enrolled in a longitudinal cohort study in Managua, Nicaragua, and attending a primary health care center from January 19, 2006, through December 31, 2023. We collected clinical records and laboratory results across the first 10 days of illness. Data were analyzed with generalized additive models, day-and-disease-specific prevalence estimates, and machine learning models.

**Findings:** We characterized 1,405 dengue, 517 chikungunya, and 522 Zika pediatric cases. We included 1,165 (47·7%) males and 1,279 (52·3%) females, with a median age of 10·0 (IQR 7·0-12·7) years. The prevalence of many clinical features exhibited by dengue, chikungunya, and Zika cases differed substantially overall, by age, and by day of illness. Dengue cases were differentiated most by abdominal pain (Prevalence difference (PD) 19·1%, 95% confidence interval (CI): 15·7%, 22·9%), leukopenia (PD 41·1%, 95% CI: 36·2%, 45·6%), nausea (PD 15·5%, 95% CI: 12·2%, 19·2%), vomiting (PD 21·9%, 95% CI: 17·9%, 26·1%), and basophilia (PD 42·3%, 95% CI: 37·4%, 47·0%); chikungunya cases were differentiated most by arthralgia (PD 60·5%, 95% CI: 56·3%, 64·2%) and the absence of leukopenia (PD −32·0%, 95% CI: −36·7%, −27·1%) and papular rash (PD −14·9%, 95% CI: −17·2%, −12·7%); and Zika cases were differentiated most by rash (PD 31·8%, 95% CI: 27·0%, 36·2%) and the lack of fever (PD −37·3%, 95% CI: −41·7%, −33·0%) and lymphocytopenia (PD −41·9%, 95% CI: −46·6%, −37·1%). Dengue and chikungunya cases exhibited similar temperature dynamics during acute illness, and their temperatures were higher than Zika cases. Sixty-two laboratory-confirmed afebrile dengue cases, which would not be captured by any widely used international case definition, presented very similarly to afebrile Zika cases, though some exhibited warning signs of disease severity. The presence of arthralgia, the presence of basophilia, and the absence of fever were the most important model-based distinguishing predictors of chikungunya, dengue, and Zika, respectively.

**Interpretations:** These findings substantially update our understanding of dengue, chikungunya, and Zika in children while identifying various clinical features that could improve differential diagnoses. The occurrence of afebrile dengue warrants reconsideration of current guidance.

**Funding:** US National Institutes of Health R01AI099631, P01AI106695, U01AI153416, U19AI118610.

**Research in context:** *Evidence before this study:* Dengue, chikungunya, and Zika co-occur in tropical and subtropical settings and cause fever, rash, and other clinical features. We reviewed international case definitions for the three diseases; the Pan American Health Organization’s (PAHO) 2022 report on their differential diagnosis; and the 80 studies underlying PAHO’s recommendations. On March 15, 2025, we queried PubMed without restrictions for “pediatric cohort” AND “dengue” AND “chikungunya” AND “Zika,” revealing that no other pediatric cohort study simultaneously reported clinical differences between the three diseases. The literature suggests that the broad and overlapping clinical features of the three diseases hamper differential diagnosis, particularly for mild forms of the diseases during acute illness, absent definitive laboratory testing. Most studies on the diseases’ clinical manifestations are in adults, and these studies constitute the main evidence base for existing case definitions. Disease presentation is more non-specific in children and adolescents than in adults, further impeding differential diagnoses in pediatric populations. Current guidelines suggest that the presence of thrombocytopenia, progressive increases in hematocrit, and leukopenia tend to distinguish dengue from chikungunya and Zika; that arthralgia is more common in chikungunya; and that pruritis is more common in Zika. All dengue case definitions in widespread use require that patients exhibit fever.

*Added value of this study:* This study follows a cohort of Nicaraguan children through multiple dengue epidemics, two large chikungunya epidemics, and one explosive Zika epidemic. Synthesizing 18 years’ worth of primary care medical records, we find clinically meaningful differences in the prevalence of many clinical features, including by day of illness and across age. In addition to verifying the clinical features PAHO identified as key distinguishing features, we also identified others, including papular rash, nausea, hemorrhagic manifestations, abdominal pain, and basophilia, that could aid differential diagnoses. As the PAHO report is based on a multitude of studies with varied age ranges, health care accessibility, overall research quality, and patient populations, this study is a complementary and important counterpart that draws from a single, well-characterized source population. Further, we identified 62 laboratory-confirmed cases of afebrile dengue (7·1% of all dengue cases since we started testing any suspected case exhibiting afebrile rash). The disease manifestations of afebrile dengue cases were generally clinically indistinguishable from afebrile Zika, although several displayed warning signs of severity. Machine learning models best distinguished chikungunya from dengue and Zika based on clinical features. Our dengue model performed well, especially in classifying febrile dengue cases. However, our Zika model struggled to properly distinguish afebrile dengue cases from afebrile Zika cases, likely due to their very similar and minimal disease presentation.

*Implications of all the available evidence:* We identify meaningful differences in the pediatric presentation and laboratory markers of dengue, chikungunya, and Zika cases that can be leveraged to improve diagnoses in the absence of laboratory testing. The distinguishing characteristics we identify could be used by future studies to build, optimize, and validate clinical prediction models. Afebrile dengue should be studied more and incorporated into future guidance, as not accounting for its existence can impede surveillance, laboratory testing strategies, clinical management, and research efforts.

## INTRODUCTION

Dengue virus (DENV), chikungunya virus (CHIKV), and Zika virus (ZIKV) are primarily transmitted by *Aedes aegypti* and *Aedes albopictus* mosquitoes. Over four billion people in the tropics and subtropics live at risk of infection (1), and climate change is exacerbating the risk for billions more (2). Where viruses overlap geographically (*e.g.*, Central and South America, Africa, Asia), explosive co-epidemics with high morbidity sometimes occur.

Four DENV serotypes (DENV-1-4) cause dengue, the world’s most common mosquito-borne disease, which ranges from undifferentiated fever to life-threatening conditions (3–5). Fever and arthralgia are especially prominent during acute-phase chikungunya, with long-term arthralgia sometimes occurring (6). Zika often presents non-specifically during childhood and as a mild, dengue-like disease in adolescence and adulthood (7). However, ZIKV infection during pregnancy can cause serious developmental complications in infants, including microcephaly.

The clinical spectrum of dengue, chikungunya, and Zika encompasses fever, rash, and other clinical features (signs, symptoms, and complete blood count results) (5). As the most common and mild forms of these typically self-limiting diseases display overlapping clinical features, primary care physicians face challenges in generating an accurate diagnosis, especially during contemporaneous outbreaks. The diseases tend to present distinctly in adults but more non-specifically in children and adolescents, posing an additional complication. Differential diagnosis and disease surveillance may thus be especially difficult in pediatric populations absent definitive laboratory testing. Further, rapid tests, which are expensive or unavailable in many resource-limited areas, may be insufficient to distinguish dengue from Zika as DENV and ZIKV are similar flaviviruses with high structural and sequence homology and antigenic cross-reactivity. In response, a recent Pan American Health Organization (PAHO) report identified clinical features useful for differential diagnosis by summarizing 80 studies with varied patient populations, age ranges, research quality, and health care settings (8). However, only one of 80 studies directly compared the prevalence of clinical features across the diseases, and that study (9) mainly focused on adults, leaving a wide gap in pediatric medicine.

We aimed to use 18 years of primary care observations from a Nicaraguan pediatric cohort to characterize distinguishing features of dengue, chikungunya, and Zika. We examined whether clinical features varied in prevalence overall, by age, by sex, and by day of illness; assessed the temporal dynamics of fever for disease-specific differences; and leveraged our study design to characterize cases (symptomatic infections) of afebrile dengue. Finally, we developed machine learning classification models to quantify how accurately differential diagnosis can be performed algorithmically given the overlapping nature of the diseases and to identify the most important clinical features for disease classification.

## METHODS

### Study design and participants

This was a single-center prospective cohort study. In 2004, the Pediatric Dengue Cohort Study (PDCS) began studying DENV infections among 2-<10-year-old children in Managua, Nicaragua (10). The PDCS expanded its eligibility criteria to include CHIKV and ZIKV before they entered the study area in August 2014 and January 2016, respectively (11,12). After several age-based expansions (2-<12 in 2008, 2-<15 in 2013), the study has included 2-<18-year-olds since 2018, after the chikungunya and Zika epidemics. The full PDCS follows ∼4,000 children who live in District II of Managua, have no plans to leave the study area within three years, attend the Health Center Sócrates Flores Vivas (HCSFV) for their medical needs, and lack immune-compromising conditions. For this study, we only included laboratory-confirmed cases of dengue, chikungunya, and Zika who were enrolled in the PDCS between January 19, 2006, and December 31, 2023, and were evaluated at the HCSFV, where PDCS participants receive free medical care 24 hours per day, 7 days per week.

Institutional Review Boards of the University of California, Berkeley, the University of Michigan, and the Nicaraguan Ministry of Health approved the PDCS protocol. Participants’ parents or legal guardians provided written informed consent. Participants ≥6 years old provided verbal assent. We involved people with lived experience in Managua, Nicaragua, across all stages of the research.

### Procedures

Participants of the PDCS are encouraged to visit the HCSFV at the first indication of any illness and to return if new signs or symptoms occur; the vast majority (95·1%) of participants comply (10). During each medical visit, information on over 100 clinical features is currently collected (appendix, p 2). Acute blood samples are collected at the first medical consult (97·0% within 1-3 days of illness) and may be collected at subsequent visits, depending on physicians’ judgement and ethical constraints on the repeated collection of blood from young children. Convalescent samples are collected 14-21 days post-illness onset (94·0% of cases).

Initially, PDCS cases suspected of dengue, chikungunya, or Zika were eligible for laboratory testing if they exhibited: 1) fever and ≥2 of the following: headache, retro-orbital pain, myalgia, arthralgia, rash, hemorrhagic manifestations, and leukopenia (1997 WHO dengue case definition (4)) or 2) undifferentiated fever without evident cause, with or without other clinical findings. After ZIKV was introduced into Managua in 2016, PDCS testing criteria expanded to encompass afebrile rash, with or without other clinical findings (7). Suspected cases were laboratory-confirmed by RT-PCR and IgM-capture and Inhibition ELISA assays using acute and convalescent samples (appendix, p 2).

Patient and laboratory data undergo extensive validation to ensure accuracy (10). For this study, we reassessed the data for consistency and completeness. Incomplete clinical records were excluded. Medical personnel reviewed moderate-to-severe discrepancies in patient data. Medically irreconcilable data were excluded.

### Statistical analysis

For this analysis, we assessed clinical features during the first 10 days of illness. Clinical features for analysis were restricted to those either 1) present in the World Health Organization (WHO) or PAHO case definitions for dengue, chikungunya, or Zika (3–5,13–15) or 2) occurring in at least 30 cases of any of the three diseases. Fever was defined as a recent history of fever or feverishness by the patient or guardian or a measured temperature of ≥38·0°C (100·4°F) during the medical consult. *Rash* denotes any type of rash.

To examine variation in the prevalence of 30 selected clinical findings, we calculated disease-specific prevalence estimates and pairwise prevalence differences across the three diseases. Prevalence-age trends (overall and stratified by sex) were estimated using logistic generalized additive models. Day-specific prevalences were also estimated. To compare the disease-specific temporal dynamics of fever, we estimated day-and-disease-specific temperature means. We precluded medication-induced bias by excluding temperature data for individuals on antipyretics when the medical consult occurred. Adapting a previously published machine learning pipeline (16,17), we developed boosted regression tree models to classify diseases based on our selected clinical findings and to identify the most important distinguishing clinical features. We trained models on individual-level data across days 1-10 of illness with each clinical feature used as a binary variable and then predicted the probability of a case being assigned to a specific disease, using misclassifications as measures of disease similarity. The appendix (pp 2-6) contains additional information on all aspects of study methodology and sensitivity analyses that 1) re-analyzed temperature dynamics using the full dataset, 2) restricted the classification models by using all 30 clinical findings across only days 1-3 of illness, and 3) restricted the classification models by using the 25 clinical findings not based on complete blood count results across days 1-10 of illness.

## RESULTS

During the study period, we confirmed 1,321 dengue, 517 chikungunya, and 522 Zika cases by molecular and serological methods (Table 1). Substantial numbers of dengue cases occurred most years, whereas chikungunya and Zika cases were concentrated in 2014-2015 and 2016, respectively (Figure S1; appendix p 17). We also detected 89 flavivirus cases (participants with DENV or ZIKV infections that could not be unambiguously distinguished by laboratory testing). Of these, 84 occurred when only DENV was circulating in Managua; we thus classified them as dengue cases. The five remaining flavivirus cases and five co-infected cases were excluded from all analyses. Among 1,405 (1,321+84) total dengue cases, 255 (18·1%) were caused by DENV-1, 425 (30·2%) by DENV-2, 306 (21·8%) by DENV-3, and 191 (13·6%) by DENV-4.

**Table 1.** Characteristics of the dengue, chikungunya, and Zika cases in the PDCS in Managua, Nicaragua, by disease (January 2006 – December 2023).

|  | **Dengue**<br>**N (%)** | **Chikungunya**<br>**N (%)** | **Zika**<br>**N (%)** |
| --- | --- | --- | --- |
| **Overall** | 1,405 (100·0) | 517 (100·0) | 522 (100·0) |
| **Epidemic years** | Most years | 2014 and 2015 | 2016 |
| **Sex** |  |  |  |
| **Male** | 666 (47·4) | 265 (51·3) | 234 (44·8) |
| **Female** | 739 (52·6) | 252 (48·7) | 288 (55·2) |
| **Age** |  |  |  |
| **Median (IQR)** | 10·5 (7·6-13·0)<sup>1</sup> | 10·2 (6·8-12·7) | 8·7 (5·8-11·5) |
| **2-<5 years** | 105 (7·5) | 70 (13·5) | 104 (19·9) |
| **5-<10 years** | 531 (37·8) | 182 (35·2) | 220 (42·1) |
| **10-<15 years** | 640 (45·6) | 265 (51·3) | 198 (37·9) |
| **15-<18 years** | 129 (9·2) | 0 (0·0) | 0 (0·0) |
| **White race** | 1,405 (100·0) | 517 (100·0) | 522 (100·0) |
| **Hispanic ethnicity** | 1,405 (100·0) | 517 (100·0) | 522 (100·0) |
| **RT-PCR confirmed** | 1,180 (84·0) | 510 (98·6) | 350 (67·0) |
| **Median day of first presentation to the health center per case (IQR)<sup>2</sup>** | 2·0 (1·0-2·0) | 2·0 (1·0-2·0) | 2·0 (1·0-2·0) |
| **Median number of medical consults per case (IQR)<sup>3</sup>** | 4·0 (2·0-6·0) | 2·0 (1·0-3·0) | 2·0 (1·0-3·0) |
| **Median number of clinical findings over days 1-10 of illness per case (IQR)<sup>4</sup>** | 8·0 (6·0-10·0) | 6·0 (5·0-8·0) | 4·0 (2·0-5·0) |
| **Median number of clinical findings over days 1-3 of illness per case (IQR)<sup>5</sup>** | 6·0 (5·0-8·0) | 6·0 (5·0-8·0) | 3·0 (2·0-5·0) |
| **Referred to pediatric hospital<sup>6</sup>** | 446 (31·7) | 42 (8·1) | 1 (0·2) |

<sup>1</sup> The statistics shown are for all dengue cases, which span an age range of 2-<18 years. The median (IQR) age for dengue cases aged 2-<15, the same age range as all chikungunya and Zika cases, is 10·0 (IQR=7·3-12·2) years.
<sup>2</sup> Chikungunya cases presented to the health center significantly earlier than dengue ( $p<0·0001$ ) and Zika cases ( $p=0·0003$ ). There was no evidence of a difference between dengue and Zika cases.
<sup>3</sup> Dengue cases had a significantly higher number of visits to the health center than chikungunya ( $p<0·0001$ ) and Zika cases ( $p<0·0001$ ). There was no evidence of a difference between chikungunya and Zika cases ( $p=0·26$ ).
<sup>4</sup> Over the first ten days of illness, dengue cases had a significantly higher number of clinical findings than chikungunya ( $p<0·0001$ ) and Zika cases ( $p<0·0001$ ). Chikungunya cases had a significantly higher number of clinical findings than Zika cases ( $p<0·0001$ ). Rash was considered collectively for this analysis; for example, if a child exhibited a rash that was both localized and macular, it was only counted once.
<sup>5</sup> Over the first three days of illness, Zika cases had a significantly fewer number of clinical findings than dengue ( $p<0·0001$ ) and chikungunya cases ( $p<0·0001$ ). There was no evidence of a difference between dengue and chikungunya cases ( $p=0·10$ ). Rash was considered collectively for this analysis.
<sup>6</sup> Dengue cases were more often referred to a pediatric tertiary referral hospital, than chikungunya ( $p<0·0001$ ) and Zika cases ( $p<0·0001$ ). Chikungunya cases were more often referred to the same hospital than Zika cases ( $p<0·0001$ ).
Abbreviations: CHIKV, chikungunya virus; DENV, dengue virus; IQR, interquartile range; PDCS, Pediatric Dengue Cohort Study; RT-PCR, reverse transcription polymerase chain reaction; SD, Standard deviation; ZIKV, Zika virus

The study sample contained 1,980 unique participants who experienced 2,444 illness episodes and had 9,087 medical consults by HCSVF physicians (Table 1). Approximately 50·0% (1,279/2,444) of all cases were female. Children 10-<15 years old constituted the largest age group. Dengue cases had significantly more clinical findings than chikungunya (p<0·0001) or Zika (p<0·0001) cases and more medical consults (p<0·0001 and p<0·0001, respectively). In addition, 31·7% (446/1,405) of dengue cases were referred to the hospital, in contrast to only 8·1% (42/517, p<0·0001) of chikungunya and 0·2% of Zika (1/522, p<0·0001) cases. Chikungunya cases reported significantly earlier to the HCSFV than dengue (p<0·0001) and Zika (p=0·0003) cases.

We first assessed the prevalence of clinical findings across diseases. Dengue cases frequently exhibited fever (95·6%, 1,343/1,405), lymphocytopenia (83·4%, 1,158/1,405), headache (75·4%, 1,060/1,405), and leukopenia (71·8%, 997/1,405) (Table 2). Fever (100·0%, 517/517), arthralgia (86·3%, 446/517), lymphocytopenia (85·6%, 441/517), and headache (76·6%, 396/517) were the most common clinical features for chikungunya patients. Rash (79·5%, 415/522), specifically generalized rash (73·9%, 386/522), was the only clinical feature occurring in ≥70·0% of Zika cases. Fever was always present for chikungunya and the vast majority of dengue cases since participants with afebrile rash were not tested until 2016.

**Table 2.** Prevalence and prevalence differences across the first ten days of illness for dengue, chikungunya, and Zika cases in the PDCS.

| Clinical finding | Dengue<br>N (%) | Chikungunya<br>N (%) | Zika<br>N (%) | Dengue vs.<br>chikungunya<br>PD (95% CI)<sup>1</sup> | Chikungunya vs.<br>Zika<br>PD (95% CI)<sup>1</sup> | Zika vs.<br>dengue<br>PD (95% CI)<sup>1</sup> |
| --- | --- | --- | --- | --- | --- | --- |
| Fever | 1,343<br>(95.6%) | 517<br>(100.0%) | 310<br>(59.4%) | -4.4%<br>(-5.6%, -3.5%) | 40.6%<br>(36.5%, 44.9%) | -36.2%<br>(-40.6%, -31.9%) |
| Rash | 531<br>(37.8%) | 281<br>(54.4%) | 415<br>(79.5%) | -16.6%<br>(-21.5%, -11.6%) | -25.1%<br>(-30.6%, -19.6%) | 41.7%<br>(37.3%, 45.8%) |
| Arthralgia | 540<br>(38.4%) | 446<br>(86.3%) | 123<br>(23.6%) | -47.8%<br>(-51.6%, -43.8%) | 62.7%<br>(57.8%, 67.2%) | -14.9%<br>(-19.2%, -10.3%) |
| Headache | 1,060<br>(75.4%) | 396<br>(76.6%) | 195<br>(37.4%) | -1.2%<br>(-5.3%, 3.3%) | 39.2%<br>(33.6%, 44.6%) | -38.1%<br>(-42.7%, -33.3%) |
| Retro-orbital pain | 378<br>(26.9%) | 67<br>(13.0%) | 37<br>(7.1%) | 13.9%<br>(10.1%, 17.5%) | 5.9%<br>(2.3%, 9.6%) | -19.8%<br>(-22.9%, -16.5%) |
| Hemorrhagic<br>manifestations | 444<br>(31.6%) | 54<br>(10.4%) | 15<br>(2.9%) | 21.2%<br>(17.4%, 24.6%) | 7.6%<br>(4.7%, 10.8%) | -28.7%<br>(-31.5%, -25.8%) |
| Myalgia | 527<br>(37.5%) | 252<br>(48.7%) | 47<br>(9.0%) | -11.2%<br>(-16.2%, -6.2%) | 39.7%<br>(34.7%, 44.6%) | -28.5%<br>(-31.9%, -24.8%) |
| Abdominal pain | 338<br>(24.1%) | 17<br>(3.3%) | 4<br>(0.8%) | 20.8%<br>(18.0%, 23.4%) | 2.5%<br>(0.9%, 4.5%) | -23.3%<br>(-25.7%, -20.9%) |
| Nausea | 309<br>(22.0%) | 24<br>(4.6%) | 9<br>(1.7%) | 17.4%<br>(14.4%, 20.1%) | 2.9%<br>(0.8%, 5.3%) | -20.3%<br>(-22.7%, -17.8%) |
| Vomiting | 391<br>(27.8%) | 49<br>(9.5%) | 13<br>(2.5%) | 18.4%<br>(14.8%, 21.7%) | 7.0%<br>(4.2%, 10.0%) | -25.3%<br>(-28.0%, -22.6%) |
| Pharyngeal<br>erythema | 646<br>(46.0%) | 195<br>(37.7%) | 108<br>(20.7%) | 8.3%<br>(3.3%, 13.1%) | 17.0%<br>(11.6%, 22.4%) | -25.3%<br>(-29.5%, -20.8%) |
| Cervical lymphadenopathy | 283<br>(20·1%) | 47<br>(9·1%) | 73<br>(14·0%) | 11·1%<br>(7·6%, 14·2%) | -4·9%<br>(-8·8%, -1·0%) | -6·2%<br>(-9·7%, -2·4%) |
| Conjunctival injection | 214<br>(15·2%) | 4<br>(0·8%) | 48<br>(9·2%) | 14·5%<br>(12·4%, 16·5%) | -8·4%<br>(-11·3%, -6·0%) | -6·0%<br>(-9·0%, -2·7%) |
| Sore throat | 272<br>(19·4%) | 28<br>(5·4%) | 46<br>(8·8%) | 13·9%<br>(11·0%, 16·7%) | -3·4%<br>(-6·6%, -0·3%) | -10·5%<br>(-13·6%, -7·2%) |
| Appetite loss | 460<br>(32·7%) | 134<br>(25·9%) | 31<br>(5·9%) | 6·8%<br>(2·2%, 11·2%) | 20·0%<br>(15·7%, 24·3%) | -26·8%<br>(-29·9%, -23·5%) |
| Cough | 289<br>(20·6%) | 53<br>(10·3%) | 41<br>(7·9%) | 10·3%<br>(6·8%, 13·6%) | 2·4%<br>(-1·1%, 6·0%) | -12·7%<br>(-15·7%, -9·4%) |
| Rhinorrhea | 276<br>(19·6%) | 44<br>(8·5%) | 32<br>(6·1%) | 11·1%<br>(7·8%, 14·2%) | 2·4%<br>(-0·8%, 5·6%) | -13·5%<br>(-16·3%, -10·4%) |
| Facial flushing | 67<br>(4·8%) | 74<br>(14·3%) | 11<br>(2·1%) | -9·5%<br>(-13·0%, -6·5%) | 12·2%<br>(9·1%, 15·7%) | -2·7%<br>(-4·2%, -0·8%) |
| Diarrhea | 76<br>(5·4%) | 11<br>(2·1%) | 8<br>(1·5%) | 3·3%<br>(1·4%, 4·9%) | 0·6%<br>(-1·1%, 2·4%) | -3·9%<br>(-5·4%, -2·1%) |
| Erythematous rash | 457<br>(32·5%) | 231<br>(44·7%) | 339<br>(64·9%) | -12·2%<br>(-17·1%, -7·2%) | -20·3%<br>(-26·1%, -14·3%) | 32·4%<br>(27·6%, 37·1%) |
| Generalized rash | 463<br>(33·0%) | 213<br>(41·2%) | 386<br>(73·9%) | -8·2%<br>(-13·2%, -3·4%) | -32·7%<br>(-38·3%, -27%) | 41·0%<br>(36·4%, 45·3%) |
| Localized rash | 119<br>(8·5%) | 99<br>(19·1%) | 43<br>(8·2%) | -10·7%<br>(-14·5%, -7·2%) | 10·9%<br>(6·8%, 15·1%) | -0·2%<br>(-2·8%, 2·8%) |
| Papular rash | 105<br>(7·5%) | 2<br>(0·4%) | 116<br>(22·2%) | 7·1%<br>(5·6%, 8·6%) | -21·8%<br>(-25·6%, -18·4%) | 14·7%<br>(11·1%, 18·7%) |
| Maculopapular rash | 71<br>(5·1%) | 1<br>(0·2%) | 102<br>(19·5%) | 4·9%<br>(3·7%, 6·2%) | -19·3%<br>(-23·0%, -16·1%) | 14·5%<br>(11·1%, 18·3%) |
| Macular rash | 176<br>(12.5%) | 72<br>(13.9%) | 182<br>(34.9%) | -1.4%<br>(-5.0%, 1.9%) | -20.9%<br>(-26.0%, -15.9%) | 22.3%<br>(18.0%, 26.8%) |
| Leukopenia | 997<br>(71.8%)<sup>2</sup> | 133<br>(25.8%)<sup>5</sup> | 215<br>(41.2%) | 46.0%<br>(41.4%, 50.3%) | -15.4%<br>(-21.0%, -9.7%) | -30.6%<br>(-35.4%, -25.7%) |
| Thrombocytopenia | 158<br>(11.4%)<sup>2</sup> | 8<br>(1.6%)<sup>5</sup> | 2<br>(0.4%) | 9.8%<br>(7.8%, 11.8%) | 1.2%<br>(0.0%, 2.7%) | -11.0%<br>(-12.8%, -9.3%) |
| Lymphocytopenia | 1,158<br>(83.4%)<sup>2</sup> | 441<br>(85.6%)<sup>5</sup> | 221<br>(42.3%) | -2.3%<br>(-5.7%, 1.5%) | 43.3%<br>(38.0%, 48.4%) | -41.0%<br>(-45.6%, -36.3%) |
| Monocytopenia | 194<br>(16.8%)<sup>3</sup> | 12<br>(2.3%)<sup>5</sup> | 7<br>(1.3%) | 14.4%<br>(11.9%, 16.9%) | 1.0%<br>(-0.7%, 2.8%) | -15.4%<br>(-17.8%, -13.0%) |
| Basophilia | 590<br>(54.9%)<sup>4</sup> | 60<br>(11.7%)<sup>5</sup> | 120<br>(23.0%) | 43.3%<br>(39.1%, 47.2%) | -11.3%<br>(-15.9%, -6.8%) | -31.9%<br>(-36.5%, -27.1%) |

<sup>1</sup> Positive differences indicate that the clinical finding was more prevalent among the first case group in the comparison, and vice versa. For example, in the column “Dengue vs. chikungunya”, fever was observed among 95.6% of the dengue cases and 100.0% of the chikungunya cases. Therefore, the prevalence difference is -4.4%.
<sup>2</sup> Percentages calculated among 1,389 (98.9%) dengue cases where leukocyte, platelet, and lymphocyte counts were available.
<sup>3</sup> Percentages calculated among 1,158 (82.4%) dengue cases where monocyte counts were available.
<sup>4</sup> Percentages calculated among 1,074 (76.4%) dengue cases where basophil counts were available.
<sup>5</sup> Percentages calculated among 515 (99.6%) chikungunya cases where complete blood count data were available.
Abbreviation: CI, confidence interval; PD, prevalence difference; PDCS, Pediatric Dengue Cohort Study

Large and significant differences in the prevalence of many clinical findings were observed across diseases (Tables 2, S1; Figure S2; appendix pp 7-9, 18). The largest differences were observed for arthralgia, which was more prevalent by 62·7 (95% CI: 57·8, 67·2) and 47·8 (95% CI: 43·8, 51·6) percentage points among chikungunya than Zika and dengue cases, respectively. The near absence of several clinical findings (*i.e.*, papular rash, conjunctival injection, abdominal pain, thrombocytopenia, and monocytopenia) for one disease but not others indicated they could be used to exclude diseases during differential diagnosis. Based on absolute and relative differences in prevalence (Table S2; appendix pp 10-12), the presence of basophilia, monocytopenia, and abdominal pain best distinguished dengue from chikungunya and Zika; the presence of arthralgia and absence of papular rash and conjunctival injection best distinguished chikungunya; and the presence of generalized, erythematous rash and the absence of fever, headache, myalgia, and lymphocytopenia best distinguished Zika.

We compared the age-varying presentation of clinical profiles that we use for laboratory testing. As age increased, a growing percentage of dengue, chikungunya, and Zika cases met the 1997 WHO dengue case definition (Figure S3; appendix p 19). Conversely, the percentage exhibiting undifferentiated fever decreased with age across diseases. Zika cases were significantly more likely to exhibit afebrile rash than dengue cases at any age.

We then examined age-prevalence trends of the underlying clinical features and found many significant differences (Figure 1). For example, the prevalence of leukopenia was >25·0 percentage points higher among dengue than Zika cases at every age, confirming earlier findings (Table 2). We also found disease-specific age patterns. Uniquely among dengue cases, the prevalence of all types of rash decreased linearly, and nausea increased linearly by age. Additionally, the prevalence of basophilia was constant and high (52·4-66·8%) across age for dengue cases, in contrast to rapid age-based waning for chikungunya and Zika cases. Among chikungunya cases, the prevalence of generalized, erythematous rash decreased from ages 2 - 10 but then rebounded and continued increasing throughout adolescence. Among Zika cases, the prevalence of fever increased with age and the prevalence of lymphocytopenia decreased before rebounding in adolescence. Age-prevalence trends did not differ by sex (Figure S4; appendix p 20).

**Figure 1.**
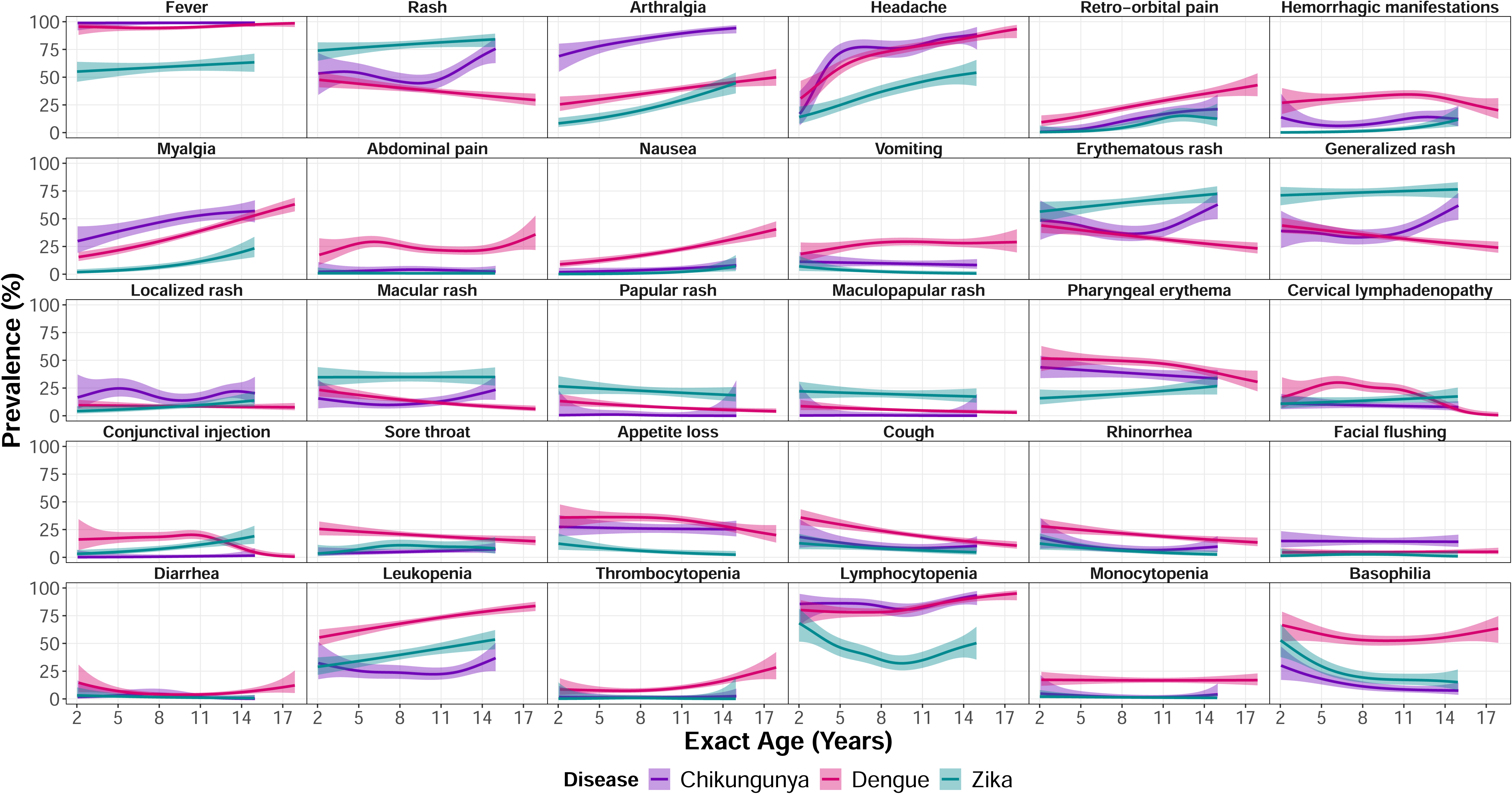
Age-prevalence trends for clinical features among the dengue, chikungunya, and Zika PDCS cases in Managua, Nicaragua (2006–2023). The solid lines indicate the point estimate of the average, and the shaded area indicates its 95% confidence band. Trends were estimated with logistic generalized additive models. A clinical finding was considered present if a case reported experiencing it during the first 10 days of illness. For this figure only, we changed the data of five chikungunya cases to not have fever in order to prevent a model convergence error, as a meaningful 95% confidence band could not be estimated in the presence of all chikungunya cases’ exhibiting fever (as was reported).

Significant differences were found after analyzing data across the first 10 days of illness (Figure 2). For example, the prevalence of fever decreased below 50·0% for Zika cases on day three (88/177, 49·7%), chikungunya cases on day four (46/122, 37·7%), and dengue cases on day six (137/555, 24·7%). Rash was most commonly observed among Zika cases, particularly on days 1-4. In contrast, among chikungunya cases, the prevalence of generalized rash increased from 11·2% (9/80) on day five to 41·5% (22/53) and on day six to 47·2% (17/36) before decreasing again; a similar pattern was observed for erythematous rash. Arthralgia was most prevalent on days 1-4 among the chikungunya cases. Compared to dengue and chikungunya cases, headache was about half as prevalent among Zika cases during days 1-2 of illness.

**Figure 2.**
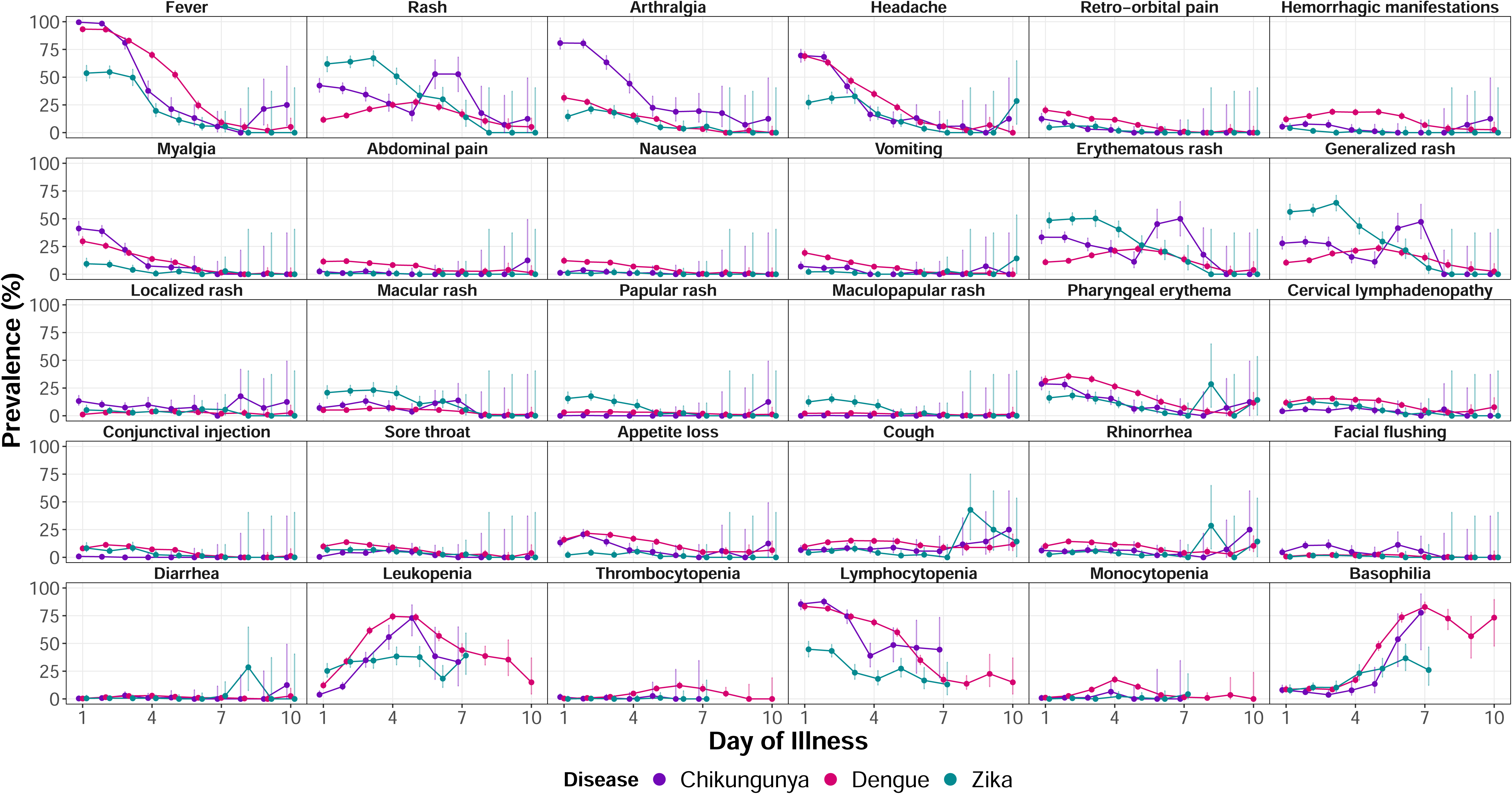
The prevalence of clinical features by disease and day of illness in the PDCS in Managua, Nicaragua (2006–2023). Dots show the day-specific average and the vertical bars correspond to its Agresti-Coull 95% confidence interval. A clinical finding was considered present if a case reported experiencing it during a given day within the first 10 days of illness. Exceedingly few chikungunya and Zika cases had blood samples taken on days 8-10 of illness. We therefore removed the unreliable point and interval estimates for leukopenia, thrombocytopenia, lymphocytopenia, monocytopenia, and basophilia for these days.

Unlike dengue and chikungunya cases, which exhibited steady increases in the prevalence of leukopenia until peaking on day five (dengue 517/702, 73·6%; chikungunya 27/37, 73·0%), the average prevalence of leukopenia was relatively stable at 32·4% (315/971 medical records) for Zika cases across days 1-7. The prevalence of basophilia was similar and low (∼10·0%) across diseases on days 1-3. Afterward, its average prevalence increased slightly to plateau at ∼27·2% (89/327 medical records) during days 4-7 of acute Zika illness. For dengue and chikungunya cases, conversely, the prevalence of basophilia substantially increased over time, exceeding 75·0% by day seven (dengue 190/229, 83·0%; chikungunya 7/9, 77·8%).

We then examined temperature readings taken at the HCSFV among cases not on antipyretic medication (n=1,833) (Table S3; appendix p 13). In general, temperature dynamics for dengue and chikungunya cases were more similar to each other than to Zika cases. Average temperatures were highest on day one across the three diseases (Figure S5; appendix p 21). While an average of 73·7% of day one temperatures for dengue (99/140) and chikungunya (66/86) cases were ≥38·0°C (100·4°F), our fever threshold, only 31·6% (12/38) of comparable Zika temperatures were ≥38·0°C. Indeed, Zika cases had mean temperatures <38·0°C across days 1-10. Only 0·1% (7/5,118) of temperature readings not influenced by antipyretics were ≥40·0°C (104·0°F), all among dengue cases.

Average fevers for dengue and chikungunya cases were significantly higher, by 0 ·5-0·9°C (0·9-1·6°F), than Zika cases during days 1-3 (dengue vs. Zika: each day p<0·0001; chikungunya vs. Zika: each day, p<0·0001). By day three, average temperatures for Zika cases returned to the interquartile range of healthy temperatures for this pediatric population (36·7-37·0°C, 98·0-98·6°F); for chikungunya and dengue cases, this occurred on days five and six, respectively. On days 4-5, fever tended to be more common among dengue than chikungunya (day four, p=0·068; day five, p=0·046) and Zika (day four p<0·0001; day five p=0·014) cases. Results were very similar between the main analyses and sensitivity analyses using the full data (Figure S6; appendix, p 22).

We identified 62 dengue cases that neither reported recent histories of fever or feverishness nor met the 38·0°C fever threshold. These cases had a median of two (IQR=1-3, range=1-9) medical consults over days 1-10 of illness. Among the 36 females and 26 males, the average age was 9·5 years (SD=3·0). These cases represent 7·1% (62/873) of dengue cases since we began testing patients exhibiting afebrile rash for DENV infection in 2016. Nine afebrile dengue cases were rRT-PCR-confirmed and caused by DENV-2. The vast majority of afebrile dengue cases (56/62, 90·3%) occurred after 2016, when ZIKV was no longer circulating in Managua. The maximum recorded temperature across the 62 cases was 37·5°C (99·5°F) (Figure S7; appendix p 23). Fifty-three (85·5%) afebrile dengue cases first reported to HCSFV within days 1-2 of illness, when temperatures for febrile dengue cases are at their highest (Figure S5; appendix p 21).

Most of the 62 afebrile dengue cases were very mild and required no hospitalization referrals (Table S4; appendix p 14-15). While none experienced severe disease, five (8·1%) exhibited dengue warning signs, as defined by the 2009 WHO guidelines (3): two with mucosal bleeding, two with abdominal pain, one with persistent vomiting. Of the 62 cases, 12 (19·4%) had no clinical findings besides rash (usually erythematous and generalized), and 17 (27·4%) exhibited only one clinical finding besides rash, predominantly (12/17, 70·6%) leukopenia. Afebrile dengue cases had significantly fewer per-case medical consults than febrile dengue cases (2·2 vs. 4·9, p<0.0001), and they most closely resembled afebrile Zika cases in terms of per-case medical consults (2·2 vs. 1·8, p=0·19), day of first medical consult (1·7 vs. 1·7, p=1·00), and number of clinical findings exhibited over the first 3 (2·5 vs. 2·3, p=0·67) and 10 days of illness (2·7 vs. 2·6, p=0·81).

Finally, we developed machine learning models that classified cases based on the 30 selected clinical findings. In general, the full models covering days 1-10 of illness had high specificity (88·8%-96·0%) and more moderate sensitivity (68·2%-86·1%) (Figure 3; Table S5, appendix p 16). The full chikungunya model correctly classified 72·5% (95% CI: 68·5, 76·2) of chikungunya cases and had a very low level (≤3·0%) of misclassification for most of the six types of non-chikungunya cases (*i.e.*, overall, febrile, and afebrile dengue and Zika cases) (Figure 3A). The full dengue model correctly classified 86·1% (95% CI: 84·0, 87·8) of all dengue cases. It correctly classified a much higher percentage of febrile (89·6%; 95% CI: 87·7, 91·2) than afebrile (15·0%; 95% CI: 7·9, 26·3) dengue cases, confirming our earlier findings that afebrile and febrile dengue present very differently. The full dengue model misclassified 18·1% (95% CI: 14·2, 22·7) of febrile Zika cases as dengue. The full Zika model correctly classified 68·2% (95% CI: 64·1, 72·1) of all Zika cases. In contrast to the dengue model, the Zika model correctly classified a higher percentage of afebrile (99·1%; 95% CI: 96·4, 100·0) than febrile (47·1%; 95% CI: 41·66, 52·7) Zika cases. Across all types of cases, the greatest percentage of misclassification was observed with the Zika model’s misclassification of 78·3% (95% CI: 66·2, 87·0) of afebrile dengue cases as Zika, confirming our earlier findings that afebrile dengue and afebrile Zika cases present very similarly. The presence of arthralgia and absence of basophilia and leukopenia most distinguished chikungunya (Figure 3B); the presence of basophilia ad leukopenia most distinguished dengue (Figure 3C); and the absence of fever most distinguished Zika (Figure 3D).

**Figure 3.**
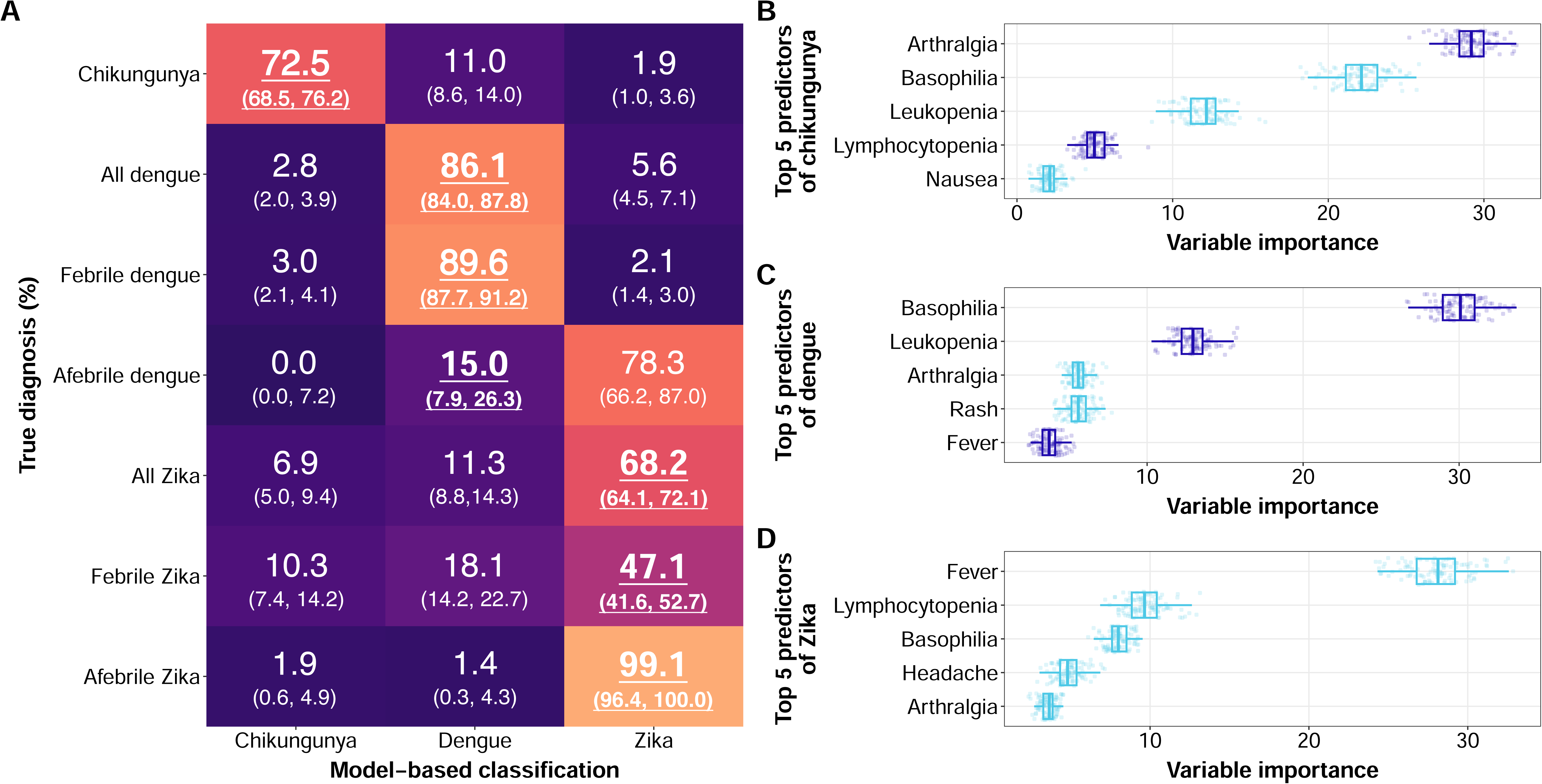
Machine learning model results for PDCS cases in Managua, Nicaragua (2006–2023). **(A)** Three separate models were constructed to classify, based on the 30 evaluated clinical findings from days 1-10 of illness, dengue from chikungunya and Zika (the dengue model), chikungunya from dengue and Zika (the chikungunya model), and Zika from dengue and chikungunya (the Zika model). The values indicate the percentage of true cases (y-axis) classified as a given disease by the disease-specific model (x-axis). For example, 72·5% of chikungunya cases were classified as chikungunya cases by the chikungunya model, and 2·8% of all dengue cases were classified as chikungunya cases by the chikungunya model. Bold and underlined percentages in panel A denote correct classifications (which an ideal classifier maximizes). Percentages in regular font represent incorrect classifications (which an ideal classifier minimizes). Parenthetical values are 95% confidence intervals. Model results are reported for all three diseases and by fever status for dengue and Zika cases. The sensitivity, specificity, positive predictive value, and negative predictive value for each model is listed in Table S5 (appendix p16). **(B-D)** The five most important clinical findings that helped correctly classify cases are shown for the chikungunya (B), dengue (C), and Zika (D) models. Dots represent variable importance values across 100 model iterations, with higher values indicating a higher importance for classification. Dark blue indicates clinical findings whose presence was important for classification; light blue indicates clinical findings whose absence was important. The top five most important variables for each model, paired with information about whether their presence or absence is relevant for classification, together determine how each given variable is helpful in classifying cases.

All three full models had increasing sensitivity with increasing pediatric age (Figure S8; appendix p 24). The chikungunya model had modest (∼50·0%) sensitivity in early childhood, likely because arthralgia, the model’s most important variable, was absent or difficult to diagnose in ∼25·0% of young children (Figure 1).

Slightly higher misclassification was observed in sensitivity analyses that included all 30 variables only through days 1-3 of illness, but results from the reduced and full models were broadly similar (Figures S9-10; appendix pp 25-26). Sensitivity analyses using data across days 1-10 of illness but excluding laboratory findings resulted in decreased sensitivity for the dengue and Zika models (Figures S11-12; appendix pp 27-28).

## DISCUSSION

To elucidate key differences in disease presentation across days 1-10 of illness, we analyzed features of dengue, chikungunya, and Zika within a single pediatric cohort. We performed the most comprehensive analysis of afebrile dengue cases to date and used machine learning to classify cases based on clinical findings. Our conclusions are strengthened by direct comparisons of clinical features across the three diseases, which only occurred in one (9) of 80 studies summarized in PAHO’s report on diagnostic guidelines (8). That one study focused mostly on adults and was ∼10-fold smaller than our current study. To our knowledge, no other pediatric cohort has simultaneously compared dengue, chikungunya, and Zika. Thus, our study meets an urgent need to identify meaningful differences across these three diseases in a well-characterized pediatric population.

PAHO’s report suggested using the occurrence of thrombocytopenia, leukopenia, and hematocrit increases to distinguish dengue; arthralgia to distinguish chikungunya; and pruritus to distinguish Zika. Our findings agree with using thrombocytopenia, leukopenia, and arthralgia. We identified several other key distinguishing features, especially at particular ages and days of illness (Table 3). Our and PAHO’s findings concur that complete blood counts are important sources of highly discriminating clinical features. In practice, physicians could compare a patient’s clinical features to our and PAHO’s lists of distinguishing characteristics to better inform differential diagnosis. This process could be automated by using computer algorithms to predict patients’ disease status based on a weighted combination of distinguishing features. While medical care for dengue, chikungunya, and Zika is strictly supportive, better-informed diagnoses would aid case management: Suspected dengue cases could receive closer monitoring and enhanced fluid management to limit severity, while suspected chikungunya cases could be advised to seek medical care if chronic polyarthralgia occurs.

**Table 3.**
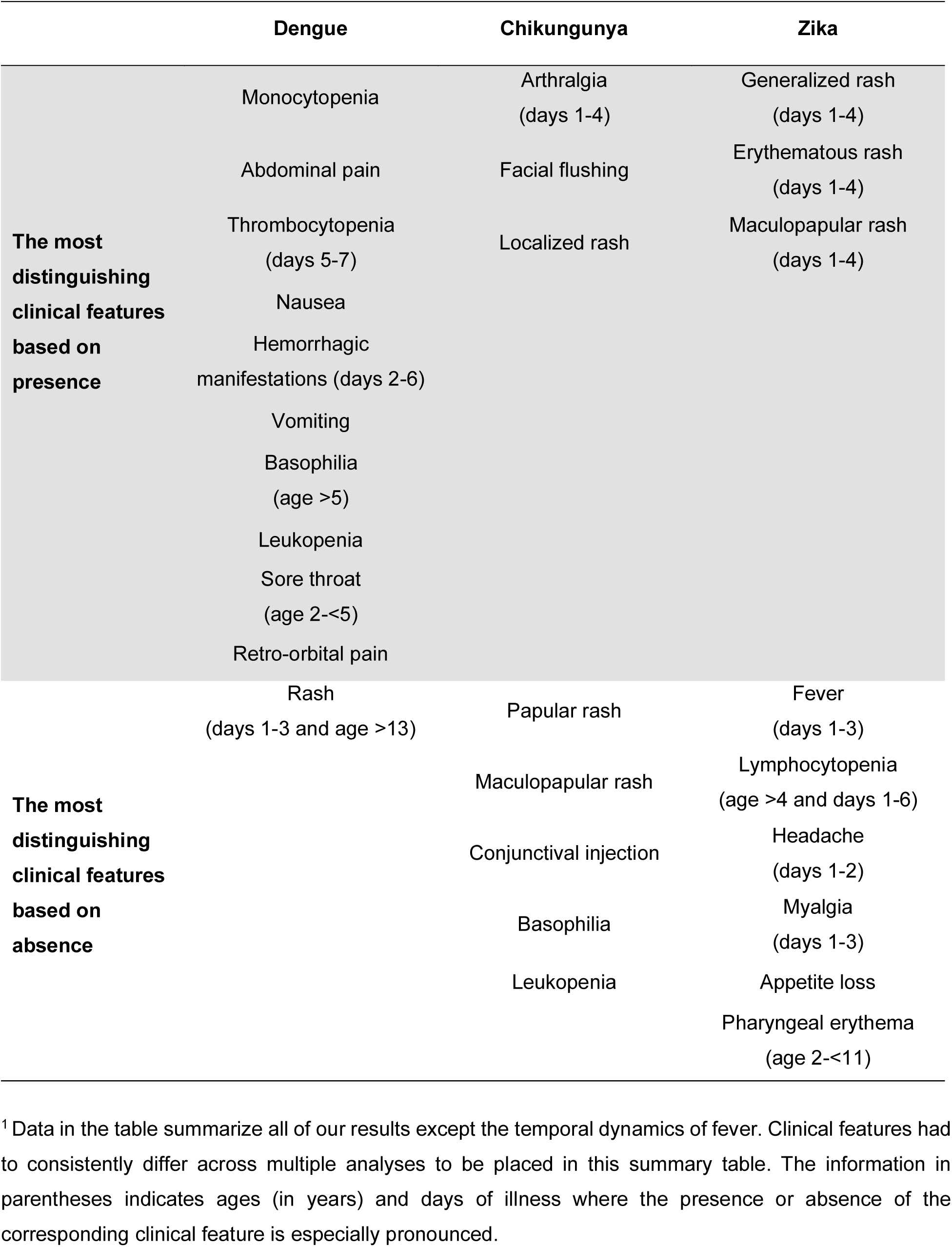
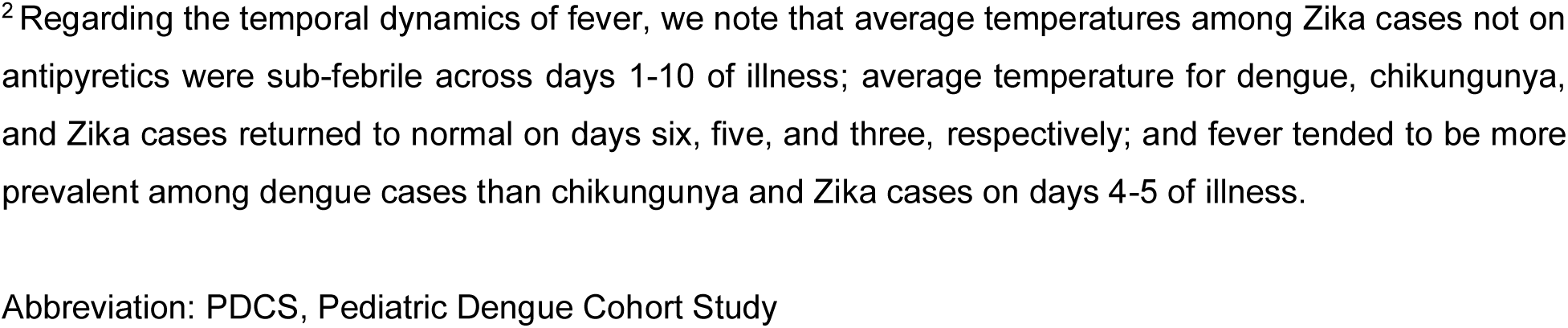
Most distinguishing clinical characteristics of dengue, chikungunya, and Zika cases in the PDCS in Managua, Nicaragua, by disease (January 2006 – December 2023).^1,2^.

|  | Dengue | Chikungunya | Zika |
| --- | --- | --- | --- |
| **The most distinguishing clinical features based on presence** | Monocytopenia | Arthralgia<br>(days 1-4) | Generalized rash<br>(days 1-4) |
|  | Abdominal pain | Facial flushing | Erythematous rash<br>(days 1-4) |
|  | Thrombocytopenia<br>(days 5-7) | Localized rash | Maculopapular rash<br>(days 1-4) |
|  | Nausea |  |  |
|  | Hemorrhagic manifestations (days 2-6) |  |  |
|  | Vomiting |  |  |
|  | Basophilia<br>(age >5) |  |  |
|  | Leukopenia |  |  |
|  | Sore throat<br>(age 2-<5) |  |  |
|  | Retro-orbital pain |  |  |
| **The most distinguishing clinical features based on absence** | Rash<br>(days 1-3 and age >13) | Papular rash | Fever<br>(days 1-3) |
|  |  | Maculopapular rash | Lymphocytopenia<br>(age >4 and days 1-6) |
|  |  | Conjunctival injection | Headache<br>(days 1-2) |
|  |  | Basophilia | Myalgia<br>(days 1-3) |
|  |  | Leukopenia | Appetite loss |
|  |  |  | Pharyngeal erythema<br>(age 2-<11) |

<sup>1</sup> Data in the table summarize all of our results except the temporal dynamics of fever. Clinical features had to consistently differ across multiple analyses to be placed in this summary table. The information in parentheses indicates ages (in years) and days of illness where the presence or absence of the corresponding clinical feature is especially pronounced.
<sup>2</sup>Regarding the temporal dynamics of fever, we note that average temperatures among Zika cases not on antipyretics were sub-febrile across days 1-10 of illness; average temperature for dengue, chikungunya, and Zika cases returned to normal on days six, five, and three, respectively; and fever tended to be more prevalent among dengue cases than chikungunya and Zika cases on days 4-5 of illness.
Abbreviation: PDCS, Pediatric Dengue Cohort Study

Our classification models algorithmically replicate the process of differential diagnosis. This allowed us to quantify the performance of modern approaches to this difficult analytical task and to identify the most important predictors of each disease. In general, our classification models performed well. However, the models’ specific weaknesses highlight that there are real limits to classification attempts of similarly presenting diseases, even under ideal conditions. These limitations are worth understanding and quantifying, since the analytic strength of modern methods cannot, in every case, overcome the irreducible overlap in clinical presentation. Nevertheless, because our approach is tailored to classification, our modeling results do robustly identify meaningfully distinguishing features of each disease.

Our chikungunya model reliably distinguished chikungunya from dengue and Zika cases, though its sensitivity diminished among young children. While the dengue model captured most febrile dengue cases, it classified 18% of febrile Zika and only 15% of afebrile dengue cases as dengue. The dengue model’s poor ability to identify afebrile dengue cases likely stemmed from their scarcity in the dataset and their non-specific and minimal clinical presentation. Interestingly, the most important predictor of dengue was basophilia, a potential biomarker for the disease. The Zika model correctly classified 99% of afebrile Zika cases, but it also misclassified 78% of afebrile dengue cases as Zika, further implying that afebrile dengue and afebrile Zika cases are only distinguishable by definitive laboratory testing. The full models and those using data only from days 1-3 of illness performed similarly, suggesting resource-limited settings experiencing concurrent epidemics should prioritize cases in early illness for laboratory testing to reduce diagnostic uncertainty and enable timelier management.

In 1952, intentional DENV infection of human volunteers (18) produced cases of afebrile dengue with rash and leukopenia, the most common presentation of our afebrile dengue cases. While our 62 afebrile dengue cases were generally mild, five exhibited warning signs. Moderate-to-severe afebrile dengue has been reported from surveillance (19) and hospital (20–23) data. The percentage of captured PDCS dengue cases that were afebrile (7·1%) is higher than other studies documenting afebrile dengue (19–26), probably because only our study systematically tested afebrile cases for many years. However, 7·1% likely represents a *lower bound* on the hidden magnitude of afebrile dengue because afebrile dengue can present without rash (23,25,26). Importantly, afebrile dengue has been observed in DENV-2 cases and in DENV-1 and DENV-4 outbreaks (24), so most, if not all, serotypes can cause afebrile dengue. The importance of afebrile dengue cases is two-fold. First, assuming an annual, conservative estimate of 51,000,000 million cases of febrile dengue (27), our conservative 7·1% estimate implies >3,897,000 afebrile cases go undetected annually. As these cases likely transmit DENV to competent vectors, surveillance, transmission, and epidemiological measures (such as R_0_) are likely systematically underestimated across epidemic settings by not considering afebrile cases. Second, weakly neutralizing anti-DENV antibodies elicited during primary infections increase the risk for severe dengue following secondary infections (28). As the immunological response to afebrile DENV infections remains uncharacterized, it is unknown if primary but afebrile dengue cases elicit severity-enhancing antibodies. Additional study is warranted.

All widely used dengue case definitions require fever (3–5). We have previously shown that the WHO 1997 and 2009 case definitions have high sensitivity but poor specificity in primary care and hospital settings (29). Assuming 7·1% of dengue cases are afebrile, loosening the fever requirement would slightly increase case definitions’ sensitivity while greatly diminishing specificity. Instead, we propose that clinical and immunological descriptions gained from additional studies of afebrile dengue be incorporated into future guidelines. For as we have previously shown for Zika (7), the spectrum of some flaviviral diseases is broader, but less febrile, than traditionally understood.

Systematic collection of medical data in a single population, large sample sizes, and modern analytic methods strengthened our study. We were limited by our testing criteria, which prevented cases of afebrile dengue and potentially afebrile chikungunya from being captured before 2016. However, our criteria reflected contemporaneous knowledge about disease presentation. Moreover, analyzing the full complexities of dengue (*e.g.,* comparisons of multiple DENV serotypes, primary vs. secondary infections, severe vs. non-severe manifestations) was beyond the scope of this study; however, these aspects are explored in depth in our companion paper focusing only on dengue cases using data from our hospital-based study and community-based cohort study over 19 years (30). Per clinical guidelines from the Nicaraguan Ministry of Health, dengue patients are referred for hospitalization at the first indication of a warning sign of disease severity. These guidelines went into effect in 2009 and precluded us from observing the full range of dengue severity in our study sample, which is based exclusively at a primary health center. Nevertheless, there was enough variation in the presentation of our dengue cases to establish how the disease tends to manifest differently than chikungunya and Zika in children. All of our participants were of similar race and ethnicity, which prevented us from examining if disease manifestations differed by racial and ethnic categories. Finally, because the HCSFV is primarily an outpatient clinic, PDCS cases are not under constant medical observation throughout the duration of their acute illness. As a result, new clinical features may have arisen during days when the cases did not report to the HCSFV for additional medical care. However, surveys of PDCS participants (10) reveal that only 4·9% of cases either consult non-PDCS medical personnel or do not consult any medical personnel when clinical features arise, indicating high participant compliance with our study protocol.

In summary, exploiting the many clinical differences we identified can enable more reliable diagnoses and better case management where confirmatory, laboratory-based diagnostic results are absent. Our afebrile dengue cases, particularly those with warning signs, emphasize that this under-characterized presentation of dengue should be incorporated into future guidance. With climate change accelerating the threat of mosquito-borne diseases (2), updated knowledge will enable improved diagnostic, surveillance, and research endeavors.

## Data Availability

Individual-level data may be shared with outside investigators following UC Berkeley IRB approval. Data will be available by request, as is required by the IRB-approved protocols for the PDCS. Please contact the UC Berkeley Center for the Protection of Human Subjects and Eva Harris to arrange for data access. Databases without names and other identifiable information were used for all analyses. Collaborating research groups and institutions will be sent coded data with all personal identifiers unlinked as well as data dictionaries and the accompanying R code.

## AUTHOR CONTRIBUTIONS

FABC and EH conceived the study. AB, AG, GK, and EH developed the study design. SO, NS, MP, AB, and GK implemented the study design and collected data. AB oversaw the laboratory testing that was performed by DC, TM, and SS. BLM, JCM, SA, LC, and ZC organized and verified the data. FBC analyzed the data and performed the statistical analyses, with CJC consulting on the machine learning models. FABC and EH drafted and revised the manuscript. All authors reviewed the manuscript and agreed to the submission of the manuscript for publication. All authors, except CJC, could access the study data if they wished. CJC provided statistical consultation without receiving the study data. The corresponding author had full access to all study data and had final responsibility in deciding to submit the paper for publication.

## ACKNOWLEDGMENTS

We are deeply grateful to our study team at the Centro de Salud Sócrates Flores Vivas, the Laboratorio Nacional de Virología at the Centro Nacional de Diagnóstico y Referencia, and the Sustainable Sciences Institute in Nicaragua. Most importantly, we sincerely thank the PDCS study participants and their families for engaging with us in the enterprise of science. This study was supported by grants R01 AI099631 (AB), P01 AI106695 (EH), U01 AI153416 (EH), and U19 AI118610 (EH) from the National Institute of Allergy and Infectious Diseases of the National Institutes of Health. Additional support for the PDCS (2004–2015) was obtained from grant VE-1 (PDVI) from the Bill and Melinda Gates Foundation as well as the FIRST grant from the Bill and Melinda Gates Foundation and the Instituto Carlos Slim de la Salud. CJC was supported by the US National Science Foundation (NSF DBI 2213854).

## DATA SHARING STATEMENT

Individual-level data may be shared with outside investigators following the approval of the UC Berkeley IRB. Data will be available by request, as is required by the IRB-approved protocols for the PDCS. Please contact the UC Berkeley Center for the Protection of Human Subjects and Eva Harris to arrange for data access. Databases without names and other identifiable information were used for all analyses. Collaborating research groups and institutions will be sent coded data with all personal identifiers unlinked as well as data dictionaries and the accompanying R code.

## REFERENCES

1. Brady OJ, Gething PW, Bhatt S, Messina JP, Brownstein JS, Hoen AG, et al. Refining the Global Spatial Limits of Dengue Virus Transmission by Evidence-Based Consensus. PLoS Negl Trop Dis. 2012 Aug;6(8):e1760--e1760.

2. Ryan SJ, Carlson CJ, Mordecai EA, Johnson LR. Global expansion and redistribution of Aedes-borne virus transmission risk with climate change. Han BA, editor. PLoS Negl Trop Dis. 2019 Mar 28;13(3):e0007213.

3. World Health Organization. Dengue: Guidelines for Diagnosis, Treatment, Prevention and Control (New Edition 2009). World Health Organization; 2009. 158 p. [cited 2025 May 10]. Available from: https://www.who.int/publications/i/item/9789241547871.

4. World Health Organization. Dengue haemorrhagic fever: Diagnosis, treatment, prevention, and control. 2nd ed. Geneva; 1997. [cited 2025 May 10]. Available from: https://iris.who.int/handle/10665/41988.

5. Pan American Health Organization. Case definitions, clinical classification, and disease phases: Dengue, Chikungunya, and Zika. Washington, DC; 2023. [cited 2025 May 10]. Available from: https://www.paho.org/en/documents/case-definitions-clinical-classification-and-disease-phases-dengue-chikungunya-and-zika.

6. Warnes CM, Andres F, Carrillo B, Zambrana JV, Mercado BL, Arguello S, et al. Longitudinal analysis of post-acute chikungunya-associated arthralgia in children and adults: A prospective cohort study in Managua, Nicaragua (2014–2018). Abu Kassim NF, editor. PLoS Negl Trop Dis. 2024 Feb 28;18(2):e0011948.

7. Burger-Calderon R, Bustos Carrillo F, Gresh L, Ojeda S, Sanchez N, Plazaola M, et al. Age-dependent manifestations and case definitions of paediatric Zika: a prospective cohort study. Lancet Infect Dis. 2020 Dec;20(3):371–80.

8. PAHO. Guidelines for the clinical diagnosis and treatment of dengue, chikungunya, and Zika. Washington, D.C.: Pan American Health Organization; 2022. [cited 2025 May 10]. Available from: https://iris.paho.org/handle/10665.2/55867.

9. Waggoner JJ, Gresh L, Vargas MJ, Ballesteros G, Tellez Y, Soda KJ, et al. Viremia and clinical presentation in Nicaraguan patients infected with Zika virus, chikungunya virus, and dengue virus. Clin Infect Dis. 2016 Dec 15;63(12):1584–90.

10. Kuan G, Gordon A, Aviles W, Ortega O, Hammond SN, Elizondo D, et al. The Nicaraguan Pediatric Dengue Cohort Study: Study design, methods, use of information technology, and extension to other infectious diseases. Am J Epidemiol. 2009 Jul 1;170(1):120–9.

11. Kuan G, Ramirez S, Gresh L, Ojeda S, Melendez M, Sanchez N, et al. Seroprevalence of anti-chikungunya virus antibodies in children and adults in Managua, Nicaragua, after the first chikungunya epidemic, 2014-2015. Bingham A, editor. PLoS Negl Trop Dis. 2016 Jun 20;10(6):e0004773.

12. Zambrana JV, Bustos Carrillo F, Burger-Calderon R, Collado D, Sanchez N, Ojeda S, et al. Seroprevalence, risk factor, and spatial analyses of Zika virus infection after the 2016 epidemic in Managua, Nicaragua. Proc Natl Acad Sci. 2018 Sep 11;115(37):9294–9.

13. World Health Organization - Regional Office for South-East Asia. Guidelines for prevention and control of Chikungunya fever. 2009. p. 19. [cited 2025 May 10]. Available from: https://www.who.int/publications/i/item/9789290223375.

14. PAHO/WHO. Zika resources: Case definitions. 2016. [cited 2025 May 10]. Available from: https://www.paho.org/en/topics/zika/zika-resources-case-definitions.

15. World Health Organization. Zika virus disease - Interim case definition. World Health Organization; 2016. [cited 2025 May 10]. Available from: https://www.who.int/publications/i/item/zika-virus-disease---interim-case-definition.

16. Becker DJ, Albery GF, Sjodin AR, Poisot T, Bergner LM, Chen B, et al. Optimising predictive models to prioritise viral discovery in zoonotic reservoirs. The Lancet Microbe. 2022 Aug 1;3(8):e625–37.

17. Mull N, Carlson CJ, Forbes KM, Becker DJ. Virus isolation data improve host predictions for New World rodent orthohantaviruses. J Anim Ecol. 2022 Jun 1;91(6):1290–302.

18. Sabin AB. Research on dengue during World War II. Am J Trop Med Hyg. 1952;1(1):30– 50.

19. Tukasan C, Furlan NB, Estofolete CF, Nogueira ML, da Silva NS. Evaluation of the importance of fever with respect to dengue prognosis according to the 2009 WHO classification: A retrospective study. BMC Infect Dis. 2017 Jan 4;17(1):1–6.

20. Amâncio FF, Heringer TP, De Oliveira CDCHB, Fassy LB, De Carvalho FB, Oliveira DP, et al. Clinical profiles and factors associated with death in adults with dengue admitted to intensive care units, Minas Gerais, Brazil. PLoS One. 2015 Jun 19;10(6):e0129046.

21. Méndez-Domínguez N, Achach-Medina K, Morales-Gual YM, Gómez-Carro S. Dengue, presentación inusual en un lactante. Reporte de un caso. Rev Chil Pediatr. 2017;88(2):275–9.

22. Badreddine S, Al-Dhaheri F, Al-Dabbagh A, Al-Amoudi A, Al-Ammari M, Elatassi N, et al. Dengue fever: Clinical features of 567 consecutive patients admitted to a tertiary care center in Saudi Arabia. Saudi Med J. 2017 Oct 1;38(10):1025–33.

23. Bhat D. The silent threats of afebrile dengue. Int J Med Sci Curr Res. 2024;7(4):449–53.

24. Yoon I-K, Rothman AL, Tannitisupawong D, Srikiatkhachorn AA, Jarman RG, Aldstadt J, et al. Underrecognized mildly symptomatic viremic dengue virus infections in rural Thai schools and villages. J Infect Dis. 2012 Aug 1;206(3):389–98.

25. Yasri S, Wiwanitkit V. Afebrile dengue myositis. Ann Trop Med Public Heal. 2016;9(5):360.

26. Knot W, Gupta N, Khatiwada S, Goyal A, Das R, Brijwal M, et al. A curious case of afebrile dengue. J Assoc Physicians India. 2018 Aug;66(8):89–90.

27. Cattarino L, Rodriguez-Barraquer I, Imai N, Cummings DAT, Ferguson NM. Mapping global variation in dengue transmission intensity. Sci Transl Med. 2020 Jan 29;12(528).

28. Katzelnick LC, Gresh L, Halloran ME, Mercado JC, Kuan G, Gordon A, et al. Antibody-dependent enhancement of severe dengue disease in humans. Science. 2017 Nov 17;358(6365):929–32.

29. Gutiérrez G, Gresh L, Pérez MÁ, Elizondo D, Avilés W, Kuan G, et al. Evaluation of the Diagnostic Utility of the Traditional and Revised WHO Dengue Case Definitions. Reithinger R, editor. PLoS Negl Trop Dis. 2013 Aug 22;7(8):e2385.

30. Narvaez F, Montenegro C, Juarez JG, Zambrana JVJV, Gonzalez K, Videa E, et al. Dengue severity by serotype and immune status in 19 years of pediatric clinical studies in Nicaragua. Marques ETA, editor. PLoS Negl Trop Dis. 2025 Jan 10;19(1):e0012811.

