## Appendix for "A comparative analysis of dengue, chikungunya, and Zika in a pediatric cohort over 18 years"

### SUPPLEMENTAL METHODS

*Study design*

The catchment area of the Health Center Sócrates Flores Visas (HCSFV), the health center where the Pediatric Dengue Cohort Study (PDCS) is based, is comprised of 18 neighborhoods in District II of Managua, Nicaragua. The population served by the study health center is approximately 62,000 persons (1). All PDCS participants receive free medical care 24 hours per day, 7 days per week at the HCSFV by study personnel. Medical data is collected when cases present to the HCSFV. As the HCSFV is a primary health care setting, it generally does not admit patients for the entire duration of their acute illness. The vast majority of participants live within the catchment area of the HCSFV. All participants are encouraged at multiple times throughout the year to report to the HCSFV at the first indication of any illness. During March and the beginning of April of each year, participants are asked to provide a healthy blood sample (2,3). During these annual samplings, demographic and household surveys are administered to PDCS participants and their family members. These surveys collect information on participants' age, sex, level of education, household characteristics, etc. Satisfaction surveys have revealed that an average of 2% of PDCS participants receive medical attention at a different medical location other than HCSFV (2). About 3% of participants report seeking no medical care during acute illnesses. Following guidelines from the Nicaraguan Ministry of Health, dengue cases with any warning sign of disease severity, following the 2009 WHO guidelines for dengue (4), are referred for hospitalization. These cases are specifically referred for follow-up at the Hospital Infantil Manuel de Jesús Rivera, a pediatric tertiary hospital in Managua, Nicaragua, that houses the Pediatric Dengue Hospital-based Study, a companion study to the PDCS.

*Laboratory methods*

Suspected dengue and chikungunya cases, caused by dengue virus (DENV) and chikungunya virus (CHIKV), respectively, were laboratory-confirmed by 1) RT-PCR of acute blood samples, with some undergoing viral isolation, 2) seroconversion by IgM capture ELISA, or 3) seroconversion or a ≥4-fold increase (DENV only) in total anti-DENV antibody titers by Inhibition ELISA (iELISA) in paired acute and convalescent samples. DENV serotyping was achieved by RT-PCR. After the introduction of Zika virus (ZIKV), a flavivirus antigenically related to DENV, acute-phase serum (or urine samples for Zika patients) of suspected cases was tested by 1) a DENV-CHIKV-ZIKV multiplex real-time RT-PCR (rRT-PCR) (5,6) or 2) a ZIKV singleplex rRT-PCR (7) alongside a pan-DENV and CHIKV rRT-PCR (8). Paired acute and convalescent serum samples from suspected Zika cases were also tested by a validated algorithm consisting of five serological assays (9,10). In practice, the iELISA mostly measures IgG anti-DENV antibodies. Additional details about the iELISA can be found in the Supplement to Katzelnick et al. (11).

*Clinical findings*

The term *clinical findings* is used to refer collectively to signs, symptoms, and complete blood count (CBC) results. This study used signs and symptoms that were captured beginning with version 4 of the PDCS medical consult form, which was implemented on January 19, 2006. Subsequent versions expanded the data capture to include additional clinical findings. Similarly, we now capture a more expanded range of complete blood count results than we did in prior years. However, signs or symptoms appearing only on subsequent forms (*e.g.,* lethargy) were not included in this report due to the high amount of missingness from cases captured using earlier versions of the form. Starting from the clinical features available in version 4 of the medical consult form and the complete blood count, we selected 25 signs and symptoms as well as 5 laboratory findings to study. We considered clinical findings that were either present in the World Health Organization and Pan American Health Organization case definitions for dengue (4,12,13), chikungunya (14), or Zika (15,16) or those presenting in at least 30 dengue, chikungunya, or Zika cases. We further ensured that selected variables had a low level of missingness. Complete blood count results that were not available for the bulk of our cases were not examined in this report. We selected the following 30 clinical findings: fever, rash (both overall and six subtypes detailed below), arthralgia, headache, retro-orbital pain, hemorrhagic manifestations (spontaneous petechiae, purpura, ecchymosis, hematoma, hemoptysis, epistaxis, gingival bleeding, melena, hematemesis, hematuria, subconjunctival hemorrhage, menorrhagia or vaginal bleeding as observed by a study physician or reported by the patient, or positive tourniquet test), myalgia, abdominal pain, nausea, vomiting, pharyngeal erythema, cervical lymphadenopathy, conjunctival injection (prominence of superficial blood vessels of the sclera), sore throat, appetite loss, coughing, rhinorrhea, facial flushing, diarrhea, leukopenia, thrombocytopenia, lymphocytopenia, monocytopenia, and basophilia.

Leukopenia was defined as <6,000 leukocytes/μl of blood for participants 2-<4 years of age, <5,000 leukocytes/μl for participants 4-<8 years of age, and <4,500 leukocytes/μl for participants ≥8 years of age. Participants with ≤100,000 platelets/µl were considered to have thrombocytopenia. Lymphocytopenia was defined as <4,000 lymphocytes/µl for participants 2-<4 years of age, <2,000 lymphocytes /µl for participants 4-<8 years of age, and <1,500 lymphocytes/µl for participants ≥8 years of age. Participants aged 2-<6 years with <300 monocytes/µl and participants aged ≥6 with <200 monocytes/µl were categorized as having monocytopenia. Participants with >100 basophils/µl were categorized as experiencing basophilia. Basophil counts were not available for cases captured in the 2023-2024 cohort year. Cases with incomplete sign and symptom data were excluded. CBC results necessitate the collection of blood samples. Because ethical considerations prevent excessive blood draws from young children, particularly across multiple medical visits on the same day, cases with missing data for CBC-derived findings were not excluded.

During the medical consultation, study physicians record five types of rash (*i.e.*, erythematous, generalized, local, macular, and papular). Participants were further categorized as having a maculopapular rash if physicians recorded both macular and papular rashes on the same medical visit. The presence of any type of rash sufficed to be categorized as having experienced rash. A rash was considered localized if it was restricted to only one of the bodily compartments (*i.e.,* head, abdomen, or extremities). A rash was considered generalized if it was present on at least two bodily compartments.

Upon intake, an HCSFV nurse takes the temperature of the PDCS participant. During the medical consultation, a HCSFV physician will retake the temperature of the PDCS participant. Both axillary temperatures are taken using a digital thermometer. For this study and because measurement error can occur with temperature readings, both temperature values taken at the medical consult were averaged. Unless noted otherwise, this average temperature is used throughout the analyses. Fever was *a priori* defined as 1) an average per-medical visit temperature ≥38·0°C (100·4°C) as measured by study staff at the HCSFV or 2) a recent history of either fever or feverishness (within a week prior to the initial medical consultation), as reported by the participant or their adult caretaker. To count as fever, the recent history of fever or feverishness reported by the participant or their caretaker did not need to include an objective measurement of the participant's temperature. When antipyretic medication use was reported by the case and their caregivers to HCSFV staff, 98·9% of the time it was acetaminophen (Figure S13). For children on antipyretics, temperatures were analyzed if medication had been taken within 30 minutes of the start of the consult (as it would be too soon for the drug to take effect) or more than four hours before the start of the consult (as the drug would have worn off by then).

In general, we consider a clinical finding to be present if it was recorded in at least one medical consultation during the first 10 days of illness, and we considered it absent if it was never reported during the same 10-day period. For analyses that focused on the per-day occurrence of clinical findings, a clinical finding was considered present if it was recorded in any of the patient's medical visits on that particular day of illness.

*Statistical and computing methods*
*Generalities*
The term *case* refers to a person with a symptomatic infection caused by either DENV, CHIKV, or ZIKV.

Afebrile dengue cases were characterized by assessing their demographics, causative DENV serotype, temperature readings, and clinical profile during their medical visits. All four DENV serotypes currently circulate in the PDCS. While serotype-specific differences in disease presentation have been previously noted (17,18), we present overall estimates of prevalence that average across the four serotypes. Continuous outcome data were compared first with Kruskal-Wallis rank sum tests and then Dunn tests with Hochberg corrections for multiple comparisons. Racial and ethnic categories used in Table 1 reflect the classifications used by the US Census Bureau.

The exposure status that defines our participants is their disease (*i.e.*, dengue, chikungunya, or Zika), and the outcomes are the 30 clinical features we assess. In the medical literature, it is extremely common to refer to the percentage of cases with a given clinical feature (even if that clinical feature is newly incident instead of already being present) as the prevalence of that clinical feature. It would be uncommon for this percentage to be referred to as the risk of the clinical feature, despite the latter being more epidemiologically precise language. As a result, we use the term *prevalence difference* to refer to the difference between two prevalence estimates. We note, though, that the epidemiological measure is formally a *risk difference*, as all participants were at risk of experiencing any clinical feature we assess. Similar logic applies to our use of the term *prevalence ratio*: Formally, the epidemiological measure under consideration is a *risk ratio*.

*Generalized additive models*

Generalized additive models (GAMs) (19,20) are semi-parametric extensions of generalized linear models (GLM). GAMs relax the linearity assumption of GLMs to account for possible non-linear trends in the data. GAMs were estimated with the mgcv R package (version 1·8-31) (20) using restricted maximum likelihood to estimate the smoothing parameters (21,22) and thin plate regression splines. The generalized Fellner-Schall optimizer (23) was only used to estimate the age-prevalence trends in Figure 1 as it generated better behaved uncertainty estimates with homogeneous data (*e.g.*, all chikungunya cases had fever). As outcome variables were binary (presence or absence of clinical findings), GAMs used binomial distributions with logit link functions.

*Prevalence differences and ratios*

Prevalence differences express the difference in prevalence for the same clinical finding across two diseases being compared. Prevalence differences across all clinical findings were estimated across the three possible pairwise comparisons of the case groups. P-values for the prevalence differences were calculated using the modern N-1 Pearson chi-square test (24) (a more accurate version of Fisher's exact test), and score-based 95% confidence intervals (25) were estimated for the prevalence differences using the epiR R package (version 1·0-13) (26).

To replicate the main analysis of the PAHO report (27) (its Annex 4, Table 1) as closely as possible using PDCS data, we analyzed prevalence differences across dengue vs. non-dengue cases (chikungunya and Zika as a composite reference group), chikungunya vs. non-chikungunya cases (dengue and Zika), and Zika vs. non-Zika cases (dengue and chikungunya). Whereas the PAHO report compares the clinical features of dengue, chikungunya, and Zika to acute febrile illnesses diseases (including malaria and leptospirosis), our analyses provide direct comparisons that isolate the prevalence of each clinical feature for each disease of interest compared to a composite reference group of the other two diseases. For these composite analyses only, we compared the 517 chikungunya cases and the 522 Zika cases to a random subset of 520 (average of 517 and 522) dengue cases. This was done so that the composite reference groups of non-chikungunya and non-Zika cases were not overly weighted towards a dengue clinical profile. We thus selected 520 dengue cases to balance the other diseases in the composite reference groups. Dengue cases for the three composite analyses were drawn from the time period starting on the date that we first tested cases presenting with afebrile rash (so that the testing criteria were the same for the Zika cases) and ending on the last date of the penultimate cohort year (as basophil counts were not available for the final cohort year). Prevalence differences, confidence intervals, and p-values were calculated as previously described. Prevalence ratios were also calculated to enable comparison using relative measures, as in the PAHO report.

*Machine learning models*

We developed machine learning models that classified disease etiology based on clinical findings, then used these models to assess 1) important clinical findings associated with each disease and 2) the level of syndromic similarity between each pair of diseases. We used a machine learning algorithm called gradient boosting, also sometimes called boosted regression trees (28), which develops an iterative set of classification trees based on the residuals of prior trees. This method allows the algorithm to focus on subsets of the data where prediction is poor, such that the model develops more 'specialized' rules that explain patterns within the data. This algorithm is therefore naturally well-suited to distinguishing diseases based on both common and rare clinical findings.

We developed models where disease identity was a binary variable, trained on individual-level clinical findings used as a set of binary covariates. We developed one model each for chikungunya, dengue, and Zika. We then applied these models to predict disease identity across all cases (including among febrile and afebrile cases of dengue and Zika) in the sample, allowing us to estimate pairwise rates of misclassification as a measure of similarity. Only cases aged 2-14 years (N=1,276 dengue, 517 chikungunya, and 522 Zika cases for both the full model using all available data for days 1-10 of illness and the reduced model using data from days 1-10 of illness but excluding complete blood count results; N=1,161 dengue, 500 chikungunya, and 500 Zika cases for the reduced model using only data from days 1-3 of illness) were input into the algorithm to enable fair comparisons among the diseases, given the identified age-related trends (Figure 1). With one minor exception, all 30 clinical features that were examined in every major analysis of this project were used in the development and characterization of each classification model. As the testing criteria for all suspected chikungunya cases required fever, we excluded fever from all chikungunya machine learning models to limit autocorrelation between predictors and the outcome variable.

Models were fit using the gbm R package (version 2·1·9) (29), using a pipeline that has previously been used to identify animal hosts of viruses based on their ecological traits (30,31). We used a fixed number of 20,000 trees to ensure convergence and selected the optimal tree depth and shrinkage parameters that maximized the area under the receiver-operator curve (AUC) of models based on a 70-30 training-test split (32). For this optimal set of hyperparameters, we generated an ensemble of 100 models with a 70-30 training-test split and used this set of models to make final predictions and estimate variable importance. The variance of variable importance values for the top five predictors across each model was visually inspected to ensure that a higher number of models was not warranted. The main models were run on the full dataset (30 clinical features across days 1-10 of illness). We thresholded model predictions to maximize the percent of training data correctly classified, using the PresenceAbsence R package (version 1·1·11) (33).

As sensitivity exercises, we developed two sets of reduced models. The first set used 30 clinical features across only days 1-3 of illness to replicate differential diagnosis under different clinical evaluation scenarios (late vs. early in disease presentation) relative to the full models. The second set of reduced models used data from days 1-10 of illness corresponding to 25 signs and symptoms, but it excluded the five complete blood count results (*i.e.*, leukopenia, thrombocytopenia, lymphocytopenia, monocytopenia, and basophilia). This second set of reduced models was useful for replicating differential diagnosis with and without access to laboratory markers relative to the full models.

Full models performed very well (AUC > 0·9) on both training data (mean model AUC for chikungunya: 0·958 (95% CI: 0·953, 0·963), dengue: 0·949 (95% CI: 0·944, 0·955), and Zika: 0·951 (95% CI: 0·943, 0·959)) and test data (mean model AUC for chikungunya: 0·943 (95% CI: 0·924, 0·959), dengue: 0·929 (95% CI: 0·915, 0·941), and Zika: 0·918 (95% CI: 0·895, 0·934)). The first set of reduced models also performed very well on both training data (mean model AUC for chikungunya: 0·952 (95% CI: 0·944, 0·960), dengue: 0·932 (95% CI: 0·922, 0·941), and Zika: 0·934 (95% CI: 0·924, 0·943)) and test data (mean model AUC for chikungunya: 0·926 (95% CI: 0·907, 0·944), dengue: 0·898 (95% CI: 0·875, 0·917), and Zika: 0·900 (95% CI: 0·876, 0·919)). The second set of reduced models performed similarly well on both training data (mean model AUC for chikungunya: 0·934 (95% CI: 0·925, 0·941), dengue: 0·890 (95% CI: 0·882, 0·897), and Zika: 0·926 (95% CI: 0·918, 0·937)) and test data (mean model AUC for chikungunya: 0·909 (95% CI: 0·890, 0·927), dengue: 0·873 (95% CI: 0·853, 0·892), and Zika: 0·895 (95% CI: 0·870, 0·914)).

*Post-hoc precision-based sample size calculations*

After using all the available data (see *Data management* section below), we used the precisely R package (version 0·1·2) (34) to conduct a comprehensive post-hoc, precision-based sample size calculation (Figure S14) following the method of Rothman and Greenland (35). In this method, the sample size is associated with the precision (width) or a 95% confidence interval rather than a postulated effect size and given level of statistical power. Our precision-based calculations are based on the width of a 95% confidence interval for the prevalence difference, since that is the main measure of association we use in the main text. Comparisons were derived for chikungunya vs. Zika and dengue vs. Zika cases; the similar numbers of chikungunya and Zika cases make a dengue vs. chikungunya analysis redundant.

Precision-based sample size calculations vary based on the difference between the prevalence of a given disease among two types of cases and the ratio of the two types of cases. Therefore, we estimated precision-based sample size calculations to account for all these factors. For each analyses we considered, 1) the ratio of the number of cases represents the actual, observed ratio; 2) three estimates of the precision-sample size association were calculated assuming the underlying prevalence of among the cases is low, medium, or high; 3) the maximum sample size of the participants in the comparison of interest was set to 10,000; and 4) the sample size was shown both for our actual study and one half as large.

*Data visualization*

Data were visualized with the ggplot2 R package (version 3·3·0) (36,37) with conditional, pointwise 95% confidence intervals (CIs) along the trendline constituting a confidence band. The patchwork R package (version 1·1·0) (38) was used to arrange distinct ggplot2 plots together to form more comprehensive figures. The R functions that generated raincloud plots were downloaded from the following website: https://gist.githubusercontent.com/benmarwick/2a1bb0133ff568cbe28d/raw/fb53bd97121f7f9ce947837ef1a4c65a73bffb3f/geom_flat_violin.R. The mean and 95% CI for day-and-disease specific temperature data was superimposed upon the half violin plot portion of the raincloud plots. Violin plots use a kernel density plot to visualize the probability distribution function of continuous data. Prevalence differences were visualized with a forest plot. Tukey-style boxplots were used to show the first, second (median), and third quartiles of continuous data. Their lower and upper whiskers extend from the first and third quartiles of the data to the smallest and largest values at most 1.5 times the interquartile range, respectively. Data outside of these whiskers are classically referred to as outliers.

*Data management*

Clinical data were extracted for all laboratory-confirmed dengue, chikungunya, and Zika cases from the first health center visit of PDCS participants from September 2, 2004, through December 31, 2023. These data included medical records extending five days before the reported illness onset date through ten days after the illness onset date, which were accessed to ensure that no relevant records were missed. In all, 10,219 medical records (one for each medical visit to the HCSFV) were extracted and assessed. Upon review, records before January 19, 2006, were dropped as several variables relevant for this study were not captured by PDCS medical consult forms (versions 1-3) used before this date. As a result, this study analyzes the 18 years' worth of primary care data from January 19, 2006, through December 31, 2023. The 10-day period of interest was indexed to the day of illness onset reported by the patient or their caretakers, not the day of fever onset, as 62 dengue cases and 212 Zika cases were afebrile throughout the duration of their acute illness and as fever was not guaranteed to occur on the first day of illness.

After reviewing the data for completeness and accuracy and dropping data before January 19, 2006, 9,114 records (89·2% of the original data) were retained and fully processed. These records corresponded to 2,454 cases, which consists of 1,321 dengue cases, 517 chikungunya cases, 522 Zika cases, 89 flaviviral cases, and 5 co-infections (1 chikungunya-Zika case, 1 dengue-chikungunya cases, and 3 dengue-Zika cases). Of the 89 flavivirus cases, 84 were reclassified as dengue cases because they occurred after the initiation of the 2017-2018 epidemic season, when ZIKV was no longer circulating. The total number of dengue cases analyzed was thus 1,405 (1,321+84). During and after the 2017-2018 epidemic season, dengue virus was the only circulating flavivirus in Managua, Nicaragua. The five flavivirus cases that occurred when both Zika virus and dengue virus were co-circulating were dropped. The five co-infections were also dropped. Of the 1,405 dengue cases, 1,180 (84·0%) were confirmed by RT-PCR; however, DENV serotype information was only available for 1,177 (99·7%) of RT-PCR-positive dengue cases.

Data management was performed in the RStudio integrated development environment (39). All data management and most analyses were performed in R version 3·6·2 (40); the machine learning models were performed in R version 4·3·2. Additional R packages contributing significantly to the data cleaning and management include readxl (version 1·3·1) (41), WriteXLS (version 5·0·0) (42), dplyr (version 1·0·3) (43), Hmisc (version 4·3-1) (44), varhandle (version 2·0·5) (45), and magrittr (version 1·5) (46).

<u>*Expanded statement of contribution from persons with lived experience*</u>

People with lived experience in Managua, Nicaragua, were involved across all stages of the research, including the funding application, study design, research implementation, and manuscript preparation. Well over half the authors (n=11, 65%) live in Managua, including the study physicians (n=4), who have decades of experience treating children with dengue, chikungunya, and Zika. Study clinicians reviewed medical records, guided the selection of clinical features to evaluate, ensured that variables were defined consistent with clinical standards set by the Nicaraguan Ministry of Health, and provided feedback on drafts of this report. The broader Nicaraguan study team includes more than 200 persons living in Managua whose professional experiences and deep knowledge of the study setting continuously shape and improve the team's research output. In all, the PDCS study team has authored over 150 research papers on the clinical, epidemiological, and immunological properties of dengue, chikungunya, and Zika viruses, all of which have featured Nicaraguan study members as authors.

### SUPPLEMENTAL TABLES

**Table S1.** Prevalence differences across dengue, chikungunya, and Zika cases in the PDCS. [1]

| Clinical finding | Dengue vs. non-dengue PD (95% CI) p-value[2] | Chikungunya vs. non-chikungunya PD (95% CI) p-value[2] | Zika vs. non-Zika PD (95% CI) p-value[2] | Dengue vs. chikungunya PD (95% CI) p-value[3] | Chikungunya vs. Zika PD (95% CI) p-value[3] | Zika vs. dengue PD (95% CI) p-value[3] |
|---|---|---|---|---|---|---|
| Fever | 13·9% (10·5%, 17·1%) <0·0001 | 23·6% (21·1%, 26·3%) <0·0001 | -37·3% (-41·7%, -33·0%) <0·0001 | -4·4% (-5·6%, -3·5%) <0·0001 | 40·6% (36·5%, 44·9%) <0·0001 | -36·2% (-40·6%, -31·9%) <0·0001 |
| Rash | -25·8% (-30·9%, -20·7%) <0·0001 | -6·0% (-11·2%, -0·8%) 0·023 | 31·8% (27·0%, 36·2%) <0·0001 | -16·6% (-21·5%, -11·6%) <0·0001 | -25·1% (-30·6%, -19·6%) <0·0001 | 41·7% (37·3%, 45·8%) <0·0001 |
| Arthralgia | -26·7% (-31·5%, -21·7%) <0·0001 | 60·5% (56·3%, 64·2%) <0·0001 | -33·5% (-38·1%, -28·7%) <0·0001 | -47·8% (-51·6%, -43·8%) <0·0001 | 62·7% (57·8%, 67·2%) <0·0001 | -14·9% (-19·2%, -10·3%) <0·0001 |
| Headache | 13·7% (8·7%, 18·5%) <0·0001 | 22·7% (17·8%, 27·3%) <0·0001 | -36·2% (-41·1%, -31·2%) <0·0001 | -1·2% (-5·3%, 3·3%) 0·60 | 39·2% (33·6%, 44·6%) <0·0001 | -38·1% (-42·7%, -33·3%) <0·0001 |
| Retro-orbital pain | 8·8% (5·1%, 12·8%) <0·0001 | 0·0% (-3·4%, 3·7%) 1·0 | -8·8% (-11·9%, -5·6%) <0·0001 | 13·9% (10·1%, 17·5%) <0·0001 | 5·9% (2·3%, 9·6%) 0·0016 | -19·8% (-22·9%, -16·5%) <0·0001 |
| Hemorrhagic manifestations | 14·3% (10·7%, 18·3%) <0·0001 | -1·5% (-4·6%, 2·0%) 0·39 | -12·8% (-15·5%, -10·2%) <0·0001 | 21·2% (17·4%, 24·6%) <0·0001 | 7·6% (4·7%, 10·8%) <0·0001 | -28·7% (-31·5%, -25·8%) <0·0001 |
| Myalgia | 0·8% (-3·9%, 5·7%) 0·73 | 29·5% (24·5%, 34·4%) <0·0001 | -30·1% (-33·9%, -26·2%) <0·0001 | -11·2% (-16·2%, -6·2%) <0·0001 | 39·7% (34·7%, 44·6%) <0·0001 | -28·5% (-31·9%, -24·8%) <0·0001 |
| Abdominal pain | 19·1% (15·7%, 22·9%) <0·0001 | -7·7% (-10·1%, -5·1%) <0·0001 | -11·5% (-13·7%, -9·4%) <0·0001 | 20·8% (18·0%, 23·4%) <0·0001 | 2·5% (0·9%, 4·5%) 0·0039 | -23·3% (-25·7%, -20·9%) <0·0001 |
| Nausea | 15·5% (12·2%, 19·2%) <0·0001 | -5·5% (-8·1%, -2·8%) 0·0002 | -9·9% (-12·2%, -7·7%) <0·0001 | 17·4% (14·4%, 20·1%) <0·0001 | 2·9% (0·8%, 5·3%) 0·0073 | -20·3% (-22·7%, -17·8%) <0·0001 |
| Vomiting | 21·9% (17·9%, 26·1%) <0·0001 | -5·7% (-8·9%, -2·2%) 0·0018 | -16·2% (-19·0%, -13·5%) <0·0001 | 18·4% (14·8%, 21·7%) <0·0001 | 7·0% (4·2%, 10·0%) <0·0001 | -25·3% (-28·0%, -22·6%) <0·0001 |
| Pharyngeal erythema | 2·6% (-2·2%, 7·5%) 0·30 | 11·5% (6·6%, 16·5%) <0·0001 | -14·0% (-18·4%, -9·4%) <0·0001 | 8·3% (3·3%, 13·1%) 0·0012 | 17·0% (11·6%, 22·4%) <0·0001 | -25·3% (-29·5%, -20·8%) <0·0001 |
| Cervical lymphadenopathy | -9·2% (-11·6%, -6·9%) <0·0001 | 0·9% (-1·9%, 4·1%) 0·53 | 8·3% (5·2%, 11·8%) <0·0001 | 11·1% (7·6%, 14·2%) <0·0001 | -4·9% (-8·8%, -1·0%) 0·014 | -6·2% (-9·7%, -2·4%) 0·0020 |
| Conjunctival injection | -2·9% (-4·7%, -0·9%) 0·0063 | -4·9% (-6·6%, -3·3%) <0·0001 | 7·7% (5·4%, 10·6%) <0·0001 | 14·5% (12·4%, 16·5%) <0·0001 | -8·4% (-11·3%, -6·0%) <0·0001 | -6·0% (-9·0%, -2·7%) 0·0006 |

| | | | | | | |
|---|---|---|---|---|---|---|
| Sore throat | 9·2%<br>(5·8%, 12·9%)<br><0·0001 | -7·2%<br>(-9·9%, -4·2%)<br><0·0001 | -2·1%<br>(-5·1%, 1·2%)<br>0·20 | 13·9%<br>(11·0%, 16·7%)<br><0·0001 | -3·4%<br>(-6·6%, -0·3%)<br>0·033 | -10·5%<br>(-13·6%, -7·2%)<br><0·0001 |
| Appetite loss | -2·8%<br>(-6·3%, 1·0%)<br>0·14 | 16·4%<br>(12·4%, 20·7%)<br><0·0001 | -13·5%<br>(-16·6%, -10·3%)<br><0·0001 | 6·8%<br>(2·2%, 11·2%)<br>0·0041 | 20·0%<br>(15·7%, 24·3%)<br><0·0001 | -26·8%<br>(-29·9%, -23·5%)<br><0·0001 |
| Cough | 7·9%<br>(4·4%, 11·7%)<br><0·0001 | -2·1%<br>(-5·3%, 1·3%)<br>0·22 | -5·7%<br>(-8·8%, -2·5%)<br>0·0009 | 10·3%<br>(6·8%, 13·6%)<br><0·0001 | 2·4%<br>(-1·1%, 6·0%)<br>0·18 | -12·7%<br>(-15·7%, -9·4%)<br><0·0001 |
| Rhinorrhea | 10·6%<br>(7·1%, 14·4%)<br><0·0001 | -3·5%<br>(-6·5%, -0·2%)<br>0·037 | -7·1%<br>(-9·9%, -4·0%)<br><0·0001 | 11·1%<br>(7·8%, 14·2%)<br><0·0001 | 2·4%<br>(-0·8%, 5·6%)<br>0·14 | -13·5%<br>(-16·3%, -10·4%)<br><0·0001 |
| Facial flushing | -6·3%<br>(-8·3%, -4·1%)<br><0·0001 | 12·3%<br>(9·4%, 15·7%)<br><0·0001 | -6·0%<br>(-8·1%, -3·8%)<br><0·0001 | -9·5%<br>(-13·0%, -6·5%)<br><0·0001 | 12·2%<br>(9·1%, 15·7%)<br><0·0001 | -2·7%<br>(-4·2%, -0·8%)<br>0·0084 |
| Diarrhea | 4·1%<br>(2·2%, 6·6%)<br><0·0001 | -1·6%<br>(-3·3%, 0·3%)<br>0·088 | -2·5%<br>(-4·1%, -0·8%)<br>0·0077 | 3·3%<br>(1·4%, 4·9%)<br>0·0021 | 0·6%<br>(-1·1%, 2·4%)<br>0·47 | -3·9%<br>(-5·4%, -2·1%)<br>0·0002 |
| Erythematous rash | -19·3%<br>(-24·3%, -14·1%)<br><0·0001 | -5·6%<br>(-10·8%, -0·3%)<br>0·037 | 24·8%<br>(19·7%, 29·8%)<br><0·0001 | -12·2%<br>(-17·1%, -7·2%)<br><0·0001 | -20·3%<br>(-26·1%, -14·3%)<br><0·0001 | 32·4%<br>(27·6%, 37·1%)<br><0·0001 |
| Generalized rash | -20·9%<br>(-26·0%, -15·7%)<br><0·0001 | -14·2%<br>(-19·3%, -8·9%)<br><0·0001 | 35·0%<br>(30·1%, 39·7%)<br><0·0001 | -8·2%<br>(-13·2%, -3·4%)<br>0·0008 | -32·7%<br>(-38·3%, -27·0%)<br><0·0001 | 41·0%<br>(36·4%, 45·3%)<br><0·0001 |
| Localized rash | -5·2%<br>(-8·3%, -1·9%)<br>0·0028 | 10·8%<br>(7·2%, 14·7%)<br><0·0001 | -5·6%<br>(-8·6%, -2·3%)<br>0·0014 | -10·7%<br>(-14·5%, -7·2%)<br><0·0001 | 10·9%<br>(6·8%, 15·1%)<br><0·0001 | -0·2%<br>(-2·8%, 2·8%)<br>0·87 |
| Papular rash | -3·1%<br>(-6·0%, 0·1%)<br>0·059 | -14·9%<br>(-17·2%, -12·7%)<br><0·0001 | 17·9%<br>(14·3%, 21·8%)<br><0·0001 | 7·1%<br>(5·6%, 8·6%)<br><0·0001 | -21·8%<br>(-25·6%, -18·4%)<br><0·0001 | 14·7%<br>(11·1%, 18·7%)<br><0·0001 |
| Maculopapular rash | -4·0%<br>(-6·6%, -1·1%)<br>0·0087 | -12·6%<br>(-14·8%, -10·6%)<br><0·0001 | 16·5%<br>(13·1%, 20·2%)<br><0·0001 | 4·9%<br>(3·7%, 6·2%)<br><0·0001 | -19·3%<br>(-23·0%, -16·1%)<br><0·0001 | 14·5%<br>(11·1%, 18·3%)<br><0·0001 |
| Macular rash | -11·0%<br>(-14·8%, -6·9%)<br><0·0001 | -10·3%<br>(-14·1%, -6·2%)<br><0·0001 | 21·2%<br>(16·6%, 25·8%)<br><0·0001 | -1·4%<br>(-5·0%, 1·9%)<br>0·42 | -20·9%<br>(-26·0%, -15·9%)<br><0·0001 | 22·3%<br>(18·0%, 26·8%)<br><0·0001 |
| Leukopenia | 41·1%<br>(36·2%, 45·6%)<br><0·0001 | -32·0%<br>(-36·7%, -27·1%)<br><0·0001 | -9·2%<br>(-14·3%, -3·9%)<br>0·0006 | 46·0%<br>(41·4%, 50·3%)<br><0·0001 | -15·4%<br>(-21·0%, -9·7%)<br><0·0001 | -30·6%<br>(-35·4%, -25·7%)<br><0·0001 |
| Thrombocytopenia | 14·2%<br>(11·3%, 17·6%)<br><0·0001 | -6·2%<br>(-8·2%, -4·2%)<br><0·0001 | -8·0%<br>(-9·9%, -6·3%)<br><0·0001 | 9·8%<br>(7·8%, 11·8%)<br><0·0001 | 1·2%<br>(0·0%, 2·7%)<br>0·054 | -11·0%<br>(-12·8%, -9·3%)<br><0·0001 |
| Lymphocytopenia | 19·0%<br>(14·6%, 23·3%)<br><0·0001 | 23·1%<br>(18·7%, 27·2%)<br><0·0001 | -41·9%<br>(-46·6%, -37·1%)<br><0·0001 | -2·3%<br>(-5·7%, 1·5%)<br>0·23 | 43·3%<br>(38·0%, 48·4%)<br><0·001 | -41·0%<br>(-45·6%, -36·3%)<br><0·0001 |
| Monocytopenia | 13·0%<br>(10·0%, 16·4%)<br><0·0001 | -5·7%<br>(-7·8%, -3·5%)<br><0·0001 | -7·3%<br>(-9·3%, -5·3%)<br><0·0001 | 14·4%<br>(11·9%, 16·9%)<br><0·0001 | 1·0%<br>(-0·7%, 2·8%)<br>0·23 | -15·4%<br>(-17·8%, -13·0%)<br><0·0001 |

| | | | | | | |
|---|---|---|---|---|---|---|
| Basophilia | 42·3%<br>(37·4%, 47·0%)<br><0·0001 | -29·6%<br>(-33·6%, -25·4%)<br><0·0001 | -12·8%<br>(-17·3%, -8·0%)<br><0·0001 | 43·3%<br>(39·1%, 47·2%)<br><0·0001 | -11·3%<br>(-15·9%, -6·8%)<br><0·0001 | -31·9%<br>(-36·5%, -27·1%)<br><0·0001 |

[1]Data in the table replicate the analysis in Annex 4 of the PAHO report (27) on the prevalence difference scale using PDCS data. Positive values indicate that the clinical feature is more prevalent in the first disease of the comparison, and negative values indicate that the clinical feature is more prevalent in the second disease or composite reference group of the comparison.

[2]For the comparisons of dengue to non-dengue (chikungunya and Zika), chikungunya to non-chikungunya (dengue and Zika), and Zika to non-Zika (dengue and chikungunya), a random subset of 520 dengue cases were selected. These analyses thus use 520 dengue cases, 517 chikungunya cases, and 522 Zika cases.

[3]These analyses use the full dataset of 1,405 dengue cases, 517 chikungunya cases, and 522 Zika cases.

Abbreviations: CI, confidence interval; PD, prevalence difference; PDCS, Pediatric Dengue Cohort Study

**Table S2.** Prevalence ratios across dengue, chikungunya, and Zika cases in the PDCS. [1]

| Clinical finding | Dengue vs. non-dengue PR (95% CI) p-value[2] | Chikungunya vs. non-chikungunya PR (95% CI) p-value[2] | Zika vs. non-Zika PR (95% CI) p-value[2] | Dengue vs. chikungunya PR (95% CI) p-value[3] | Chikungunya vs. Zika PR (95% CI) p-value[3] | Zika vs. dengue PR (95% CI) p-value[3] |
|---|---|---|---|---|---|---|
| Fever | 1·2 (1·1, 1·2) <0·0001 | 1·3 (1·2, 1·4) <0·0001 | 0·6 (0·6, 0·7) <0·0001 | 1·0 (0·9, 1·1) <0·0001 | 1·7 (1·5, 1·8) <0·0001 | 0·6 (0·6, 0·7) <0·0001 |
| Rash | 0·6 (0·5, 0·7) <0·0001 | 0·9 (0·8, 1·0) 0·023 | 1·7 (1·5, 1·8) <0·0001 | 0·7 (0·6, 0·8) <0·0001 | 0·7 (0·6, 0·7) <0·0001 | 2·1 (1·9, 2·3) <0·0001 |
| Arthralgia | 0·5 (0·4, 0·6) <0·0001 | 3·3 (3·0, 3·7) <0·0001 | 0·4 (0·3, 0·5) <0·0001 | 0·4 (0·4, 0·5) <0·0001 | 3·7 (3·1, 4·3) <0·0001 | 0·6 (0·5, 0·7) <0·0001 |
| Headache | 1·2 (1·1, 1·3) <0·0001 | 1·4 (1·3, 1·5) <0·0001 | 0·5 (0·5, 0·6) <0·0001 | 1·0 (0·9, 1·0) 0·60 | 2·1 (1·8, 2·3) <0·0001 | 0·5 (0·4, 0·6) <0·0001 |
| Retro-orbital pain | 1·9 (1·5, 2·4) <0·0001 | 1·0 (0·8, 1·3) 1·0 | 0·4 (0·3, 0·6) <0·0001 | 2·1 (1·6, 2·6) <0·0001 | 1·8 (1·3, 2·7) 0·0016 | 0·3 (0·2, 0·4) <0·0001 |
| Hemorrhagic manifestations | 3·2 (2·4, 4·2) <0·0001 | 0·9 (0·6, 1·2) 0·39 | 0·2 (0·1, 0·3) <0·0001 | 3·0 (2·3, 3·9) <0·0001 | 3·6 (2·1, 6·3) <0·0001 | 0·1 (0·1, 0·1) <0·0001 |
| Myalgia | 1·0 (0·9, 1·2) 0·73 | 2·5 (2·2, 2·9) <0·0001 | 0·2 (0·2, 0·3) <0·0001 | 0·8 (0·7, 0·9) <0·0001 | 5·4 (4·1, 7·2) <0·0001 | 0·2 (0·2, 0·3) <0·0001 |
| Abdominal pain | 10·5 (6·7, 16·4) <0·0001 | 0·3 (0·2, 0·5) <0·0001 | 0·1 (0·0, 0·2) <0·0001 | 7·3 (4·6, 11·8) <0·0001 | 4·3 (1·5, 12·1) 0·0039 | 0·0 (0·0, 0·1) <0·0001 |
| Nausea | 5·9 (4, 8·6) <0·0001 | 0·5 (0·3, 0·7) 0·0002 | 0·1 (0·1, 0·3) <0·0001 | 4·7 (3·2, 7·1) <0·0001 | 2·7 (1·3, 5·6) 0·0073 | 0·1 (0·0, 0·1) <0·0001 |
| Vomiting | 4·7 (3·5, 6·2) <0·0001 | 0·6 (0·5, 0·8) 0·0018 | 0·1 (0·1, 0·2) <0·0001 | 2·9 (2·2, 3·9) <0·0001 | 3·8 (2·1, 6·9) <0·0001 | 0·1 (0·1, 0·2) <0·0001 |
| Pharyngeal erythema | 1·1 (0·9, 1·3) 0·30 | 1·4 (1·2, 1·7) <0·0001 | 0·6 (0·5, 0·7) <0·0001 | 1·2 (1·1, 1·4) 0·0012 | 1·8 (1·5, 2·2) <0·0001 | 0·4 (0·4, 0·5) <0·0001 |
| Cervical lymphadenopathy | 0·2 (0·1, 0·4) <0·0001 | 1·1 (0·8, 1·6) 0·53 | 2·5 (1·8, 3·4) <0·0001 | 2·2 (1·7, 3·0) <0·0001 | 0·7 (0·5, 0·9) 0·014 | 0·7 (0·5, 0·9) 0·0020 |
| Conjunctival injection | 0·4 (0·2, 0·8) 0·0063 | 0·1 (0·1, 0·4) <0·0001 | 6·4 (3·6, 11·2) <0·0001 | 19·7 (7·7, 50·9) <0·0001 | 0·1 (0·0, 0·2) <0·0001 | 0·6 (0·4, 0·8) 0·0006 |
| Sore throat | 2·3 (1·7, 3·1) <0·0001 | 0·4 (0·3, 0·6) <0·0001 | 0·8 (0·6, 1·1) 0·20 | 3·6 (2·5, 5·2) <0·0001 | 0·6 (0·4, 1·0) 0·033 | 0·5 (0·3, 0·6) <0·0001 |

| | | | | | | |
|---|---|---|---|---|---|---|
| Appetite loss | 0·8<br>(0·6, 1·1)<br>0·14 | 2·7<br>(2·2, 3·5)<br><0·0001 | 0·3<br>(0·2, 0·4)<br><0·0001 | 1·3<br>(1·1, 1·5)<br>0·0041 | 4·4<br>(3·0, 6·3)<br><0·0001 | 0·2<br>(0·1, 0·3)<br><0·0001 |
| Cough | 1·9<br>(1·4, 2·4)<br><0·0001 | 0·8<br>(0·6, 1·1)<br>0·22 | 0·6<br>(0·4, 0·8)<br>0·0009 | 2·0<br>(1·5, 2·6)<br><0·0001 | 1·3<br>(0·9, 1·9)<br>0·18 | 0·4<br>(0·3, 0·5)<br><0·0001 |
| Rhinorrhea | 2·4<br>(1·8, 3·2)<br><0·0001 | 0·7<br>(0·5, 1·0)<br>0·037 | 0·5<br>(0·3, 0·7)<br><0·0001 | 2·3<br>(1·7, 3·1)<br><0·0001 | 1·4<br>(0·9, 2·1)<br>0·14 | 0·3<br>(0·2, 0·4)<br><0·0001 |
| Facial flushing | 0·2<br>(0·1, 0·4)<br><0·0001 | 7·1<br>(4·4, 11·4)<br><0·0001 | 0·3<br>(0·1, 0·5)<br><0·0001 | 0·3<br>(0·2, 0·5)<br><0·0001 | 6·8<br>(3·7, 12·6)<br><0·0001 | 0·4<br>(0·2, 0·8)<br>0·0084 |
| Diarrhea | 3·3<br>(1·9, 5·7)<br><0·0001 | 0·6<br>(0·3, 1·1)<br>0·088 | 0·4<br>(0·2, 0·8)<br>0·0077 | 2·5<br>(1·4, 4·7)<br>0·0021 | 1·4<br>(0·6, 3·3)<br>0·47 | 0·3<br>(0·1, 0·6)<br>0·0002 |
| Erythematous rash | 0·6<br>(0·6, 0·7)<br><0·0001 | 0·9<br>(0·8, 1·0)<br>0·037 | 1·6<br>(1·5, 1·8)<br><0·0001 | 0·7<br>(0·6, 0·8)<br><0·0001 | 0·7<br>(0·6, 0·8)<br><0·0001 | 2·0<br>(1·8, 2·2)<br><0·0001 |
| Generalized rash | 0·6<br>(0·6, 0·7)<br><0·0001 | 0·7<br>(0·7, 0·8)<br><0·0001 | 1·9<br>(1·7, 2·1)<br><0·0001 | 0·8<br>(0·7, 0·9)<br>0·0008 | 0·6<br>(0·5, 0·6)<br><0·0001 | 2·2<br>(2·0, 2·5)<br><0·0001 |
| Localized rash | 0·6<br>(0·4, 0·9)<br>0·0028 | 2·3<br>(1·8, 3·0)<br><0·0001 | 0·6<br>(0·4, 0·8)<br>0·0014 | 0·4<br>(0·3, 0·6)<br><0·0001 | 2·3<br>(1·7, 3·3)<br><0·0001 | 1·0<br>(0·7, 1·4)<br>0·87 |
| Papular rash | 0·7<br>(0·5, 1·0)<br>0·059 | 0·0<br>(0·0, 0·1)<br><0·0001 | 5·1<br>(3·7, 7·1)<br><0·0001 | 19·3<br>(5·3, 71·1)<br><0·0001 | 0·0<br>(0·0, 0·1)<br><0·0001 | 3·0<br>(2·3, 3·8)<br><0·0001 |
| Maculopapular rash | 0·6<br>(0·4, 0·9)<br>0·0087 | 0·0<br>(0·0, 0·1)<br><0·0001 | 6·3<br>(4·3, 9·3)<br><0·0001 | 26·1<br>(4·6, 149·4)<br><0·0001 | 0·0<br>(0·0, 0·1)<br><0·0001 | 3·9<br>(2·9, 5·1)<br><0·0001 |
| Macular rash | 0·6<br>(0·4, 0·7)<br><0·0001 | 0·6<br>(0·5, 0·7)<br><0·0001 | 2·5<br>(2·1, 3·1)<br><0·0001 | 0·9<br>(0·7, 1·2)<br>0·42 | 0·4<br>(0·3, 0·5)<br><0·0001 | 2·8<br>(2·3, 3·3)<br><0·0001 |
| Leukopenia | 2·2<br>(2·0, 2·5)<br><0·0001 | 0·4<br>(0·4, 0·5)<br><0·0001 | 0·8<br>(0·7, 0·9)<br>0·0006 | 2·8<br>(2·4, 3·2)<br><0·0001 | 0·6<br>(0·5, 0·7)<br><0·0001 | 0·6<br>(0·5, 0·6)<br><0·0001 |
| Thrombocytopenia | 15·8<br>(8·3, 29·9)<br><0·0001 | 0·2<br>(0·1, 0·4)<br><0·0001 | 0·0<br>(0·0, 0·2)<br><0·0001 | 7·3<br>(3·7, 14·6)<br><0·0001 | 4·1<br>(1·0, 16·8)<br>0·054 | 0·0<br>(0·0, 0·1)<br><0·0001 |
| Lymphocytopenia | 1·3<br>(1·2, 1·4)<br><0·0001 | 1·4<br>(1·3, 1·5)<br><0·0001 | 0·5<br>(0·5, 0·6)<br><0·0001 | 1·0<br>(0·9, 1·0)<br>0·23 | 2·0<br>(1·8, 2·3)<br><0·0001 | 0·5<br>(0·5, 0·6)<br><0·0001 |
| Monocytopenia | 8·1<br>(5·0, 13·2)<br><0·0001 | 0·3<br>(0·2, 0·5)<br><0·0001 | 0·2<br>(0·1, 0·3)<br><0·0001 | 7·2<br>(4·1, 12·7)<br><0·0001 | 1·7<br>(0·7, 4·3)<br>0·23 | 0·1<br>(0·0, 0·2)<br><0·0001 |
| Basophilia | 3·4<br>(3·0, 4·0)<br><0·0001 | 0·3<br>(0·2, 0·4)<br><0·0001 | 0·6<br>(0·5, 0·8)<br><0·0001 | 4·7<br>(3·7, 6·0)<br><0·0001 | 0·5<br>(0·4, 0·7)<br><0·0001 | 0·4<br>(0·4, 0·5)<br><0·0001 |

[1] Data in the table replicate the analysis in Annex 4 of the PAHO report (27) on the prevalence ratio scale using PDCS data. Values above 1 indicate that the clinical feature is more prevalent in the first disease of the comparison, and values below 1 indicate that the clinical feature is more prevalent in the second disease or

composite reference group of the comparison. The p-values are the same as in Table S1 since a test of whether the prevalence difference is 0 is statistically equivalent to testing whether the prevalence ratio is 1.

[2] For the comparisons of dengue to non-dengue (chikungunya and Zika), chikungunya to non-chikungunya (dengue and Zika), and Zika to non-Zika (dengue and chikungunya), a random subset of 520 dengue cases were selected alongside all 517 chikungunya cases and 522 Zika cases.

[3] These analyses use the full dataset of 1,405 dengue cases, 517 chikungunya cases, and 522 Zika cases.

Abbreviations: CI, confidence interval; PD, prevalence difference; PDCS, Pediatric Dengue Cohort Study

**Table S3.** Fever and use of antipyretic medication by day of illness at medical consults in the PDCS.

| | Dengue N (%) | Chikungunya N (%) | Zika N (%) |
|---|---|---|---|
| **Number of medical consults** | 6,722 (100·0) | 1,215 (100·0) | 1,150 (100·0) |
| **Experienced fever[1]** | | | |
| **Day 1** | 494 (93.4) | 245 (99.2) | 106 (54.1) |
| **Day 2** | 1,004 (90.5) | 385 (98.5) | 178 (54.9) |
| **Day 3** | 925 (77.9) | 191 (80.9) | 90 (47.9) |
| **Day 4** | 747 (64.4) | 47 (36.4) | 34 (19.4) |
| **Day 5** | 482 (45.4) | 17 (20.5) | 14 (11.5) |
| **Day 6** | 172 (22.0) | 8 (14.8) | 5 (5.7) |
| **Day 7** | 45 (9.6) | 2 (5.6) | 2 (5.6) |
| **Day 8** | 13 (5.3) | 0 (0.0) | 0 (0.0) |
| **Day 9** | 3 (2.9) | 3 (21.4) | 0 (0.0) |
| **Day 10** | 4 (5.2) | 2 (25.0) | 0 (0.0) |
| **Not under the influence of an anti-pyretic[1,2]** | | | |
| **Day 1** | 87 (35.2) | 141 (26.7) | 38 (19.4) |
| **Day 2** | 143 (36.6) | 423 (38.1) | 101 (31.2) |
| **Day 3** | 133 (56.4) | 550 (46.3) | 105 (55.9) |
| **Day 4** | 96 (74.4) | 616 (53.1) | 118 (67.4) |
| **Day 5** | 80 (96.4) | 739 (69.7) | 79 (64.8) |
| **Day 6** | 47 (87.0) | 638 (81.5) | 58 (66.7) |
| **Day 7** | 34 (94.4) | 415 (88.7) | 28 (77.8) |
| **Day 8** | 17 (100.0) | 231 (94.7) | 3 (42.9) |
| **Day 9** | 13 (92.9) | 96 (93.2) | 5 (62.5) |
| **Day 10** | 7 (87.5) | 72 (93.5) | 5 (71.4) |

[1] Percentages are calculated using the number of disease- and day-specific medical records as the denominator.

[2] As in the analysis of temperatures for children not under the effect of anti-pyretic medication, we define not under the influence of antipyretic medication as participants reporting 1) not taking any antipyretic, 2) taking antipyretic medication within 30 minutes of the start of the consult (as it would be too soon for the drug to take effect), or 3) taking antipyretic medication more than four hours before the start of the consult (as the drug would have worn off by then).

**Table S4.** Clinical findings of the 62 afebrile dengue cases identified in the PDCS in Managua, Nicaragua, from July 2016, when the PDCS started testing cases with afebrile rash, through December 2023.

| Clinical finding | N (%) |
|---|---|
| Fever | 0 (0·0) |
| Rash[1] | 62 (100·0) |
| Arthralgia | 1 (1·6) |
| Headache | 14 (22·6) |
| Retro-orbital pain | 2 (3·2) |
| Hemorrhagic manifestations[2] | 2 (3·2) |
| Myalgia | 1 (1·6) |
| Abdominal pain | 2 (3·2) |
| Nausea | 1 (1·6) |
| Vomiting | 1 (1·6) |
| Pharyngeal erythema | 6 (9·7) |
| Cervical lymphadenopathy | 0 (0·0) |
| Conjunctival injection | 1 (1·6) |
| Sore throat | 2 (3·2) |
| Appetite loss | 2 (3·2) |
| Cough | 3 (4·8) |
| Rhinorrhea | 4 (6·5) |
| Facial flushing | 1 (1·6) |
| Diarrhea | 0 (0·0) |
| Erythematous rash | 55 (88·7) |
| Generalized rash | 61 (98·4) |
| Localized rash | 3 (4·8) |
| Papular rash | 27 (43·5) |
| Maculopapular rash[3] | 23 (37·1) |

| | |
|---|---|
| Macular rash | 33 (53·2) |
| Leukopenia | 33 (53·2) |
| Thrombocytopenia | 0 (0·0) |
| Lymphocytopenia | 17 (27·4) |
| Monocytopenia | 1 (1·6) |
| Basophilia[4] | 11 (17·7) |

[1] Rash refers to any kind of rash.

[2] Hemorrhagic manifestations were defined as spontaneous petechiae, purpura, ecchymosis, hematoma, hemoptysis, epistaxis, gingival bleeding, melena, hematemesis, hematuria, subconjunctival hemorrhage, menorrhagia or vaginal bleeding as observed by a study physician or reported by the patient, or positive tourniquet test.

[3] Maculopapular rash was defined as a rash that was both macular and papular during a single medical visit.

[4] Percentage calculated among 59 (95·2%) afebrile Zika cases where basophil counts were available.

Abbreviations: PDCS, Pediatric Dengue Cohort Study

**Table S5.** Sensitivity, specificity, positive predictive value, and negative predictive value for each of the classification models.

| | **Chikungunya model** Percent (95% CI) | **Dengue model** Percent (95% CI) | **Zika model** Percent (95% CI) |
|---|---|---|---|
| **Full model: All clinical features, days 1-10 of illness** | | | |
| **Sensitivity (%)** | 72·5 (68·5, 76·2) | 86·1 (84·0, 87·8) | 68·2 (64·1, 72·1) |
| **Specificity (%)** | 96·0 (95·0, 96·8) | 88·8 (86·8, 90·6) | 95·4 (94·4, 96·3) |
| **Positive predictive value (%)** | 83·9 (80·2, 87·0) | 90·4 (88·7, 92·0) | 81·3 (77·3, 84·7) |
| **Negative predictive value (%)** | 92·4 (91·1, 93·5) | 83·8 (81·5, 85·9) | 91·2 (89·8, 92·4) |
| **Reduced model: All clinical features, days 1-3 of illness** | | | |
| **Sensitivity (%)** | 67·4 (63·2, 71·4) | 84·3 (82·1, 86·3) | 70·4 (66·3, 74·2) |
| **Specificity (%)** | 96·6 (95·6, 97·3) | 85·0 (82·6, 87·1) | 94·0 (92·7, 95·0) |
| **Positive predictive value (%)** | 85·5 (81·7, 88·7) | 86·7 (84·6, 88·6) | 77·9 (73·8, 81·5) |
| **Negative predictive value (%)** | 90·8 (89·3, 92·0) | 82·4 (79·9, 84·6) | 91·3 (89·9, 92·6) |
| **Reduced model: All non-laboratory clinical features, days 1-10 of illness** | | | |
| **Sensitivity (%)** | 76·8 (73·0, 80·2) | 76·8 (74·4, 79·0) | 56·5 (52·2, 60·7) |
| **Specificity (%)** | 91·2 (89·8, 92·4) | 85·7 (83·4, 87·7) | 96·7 (95·8, 97·4) |
| **Positive predictive value (%)** | 71·4 (67·5, 75·0) | 86·8 (84·7, 88·7) | 83·3 (79·1, 86·9) |
| **Negative predictive value (%)** | 93·2 (91·9, 94·3) | 75·0 (72·5, 77·4) | 88·4 (86·9, 89·8) |

**SUPPLEMENTAL FIGURES**

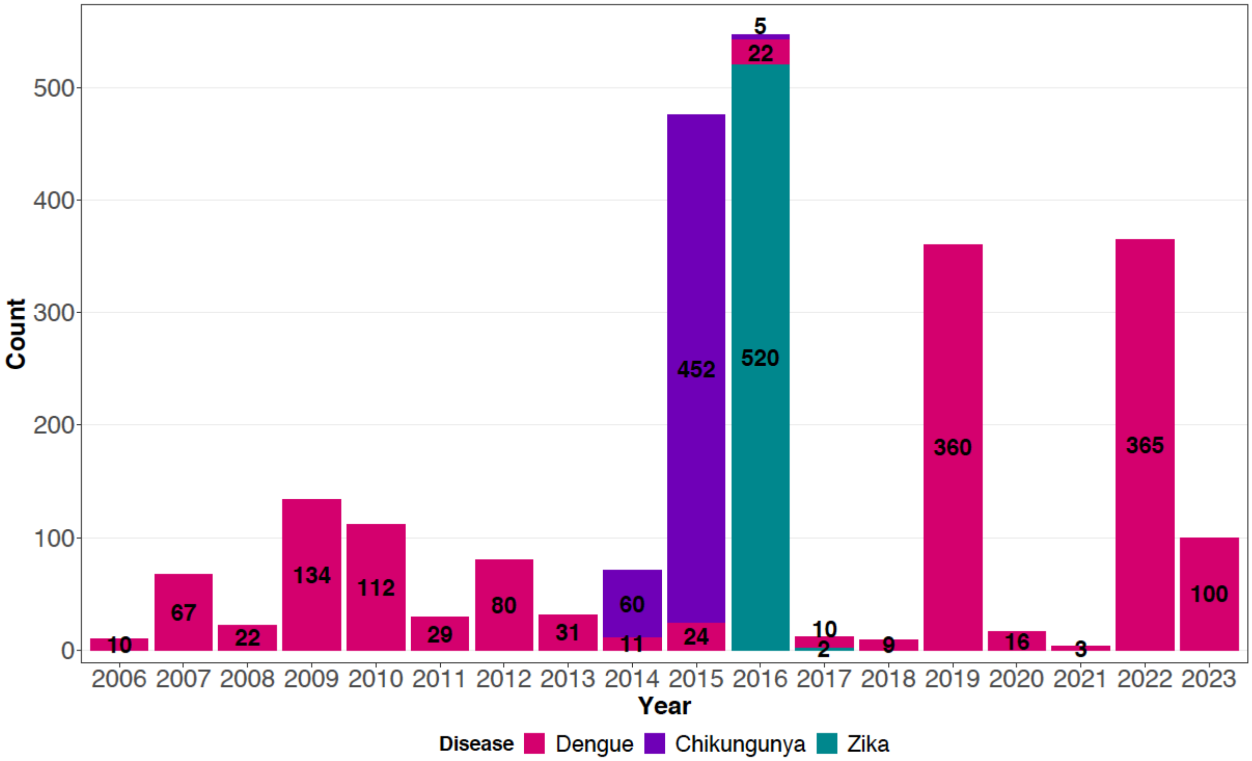

**Figure S1. Dengue, chikungunya, and Zika case counts in the PDCS by calendar year.** The count for each type of case is displayed at the midpoint of each stacked bar. The case count covers the period from January 19, 2006, through December 31, 2023.

Abbreviations: PDCS, Pediatric Dengue Cohort Study.

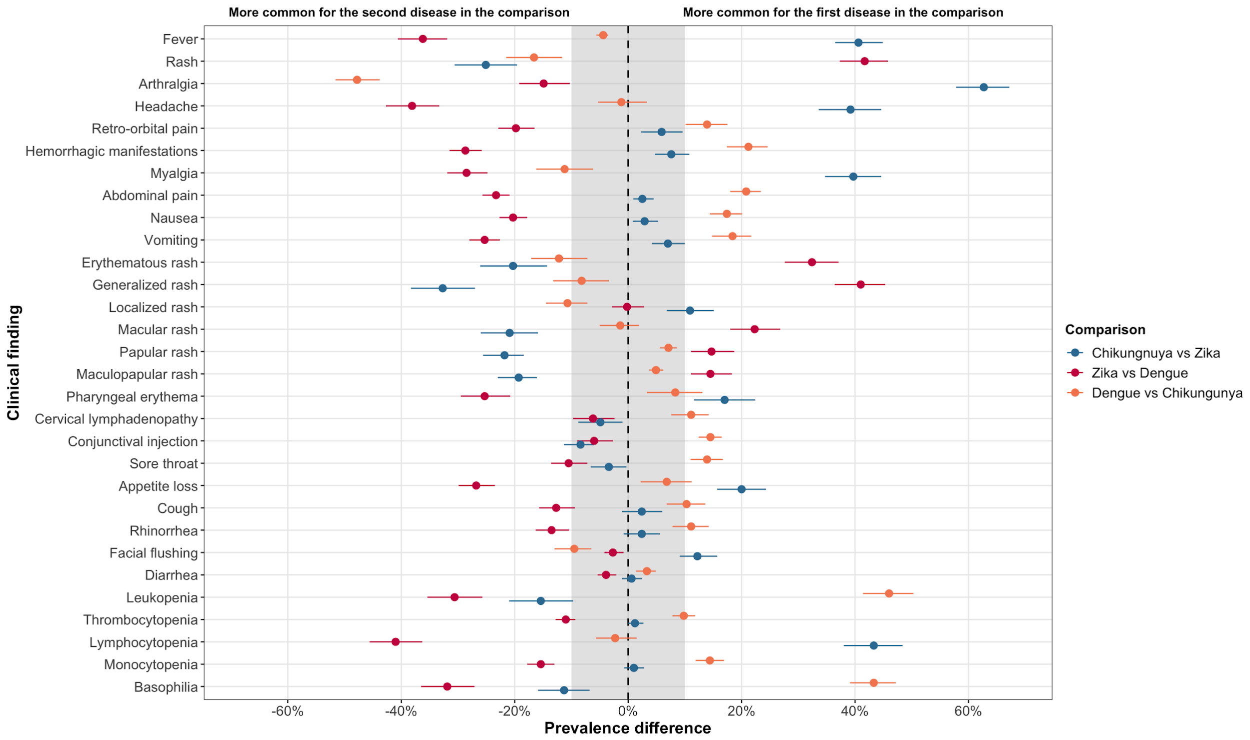

**Figure S2. Prevalence differences for each clinical finding among all pairwise comparisons of dengue, chikungunya, and Zika PDCS cases in Managua, Nicaragua (2006-2023).** This figure depicts the data in the three rightmost columns in Table 2. Positive differences (those to the right of the dashed line) indicate that the clinical finding is more prevalent among the first case group in the comparison, and vice versa. For example, fever was observed among 100% of the chikungunya cases and 60% of the Zika cases. Therefore, the prevalence difference is +40%, as shown on the right side of the figure in blue. Dots indicate the point estimate of the average and the length of the bar indicates its 95% confidence interval. Intervals that include the null value of 0 are not statistically significant at the alpha=0·05 level; intervals that exclude the null correspond to statistically significant differences. The arbitrary range of -10% to +10% is shaded light gray to indicate small prevalence differences, regardless of statistical significance, that may not be clinically important. A clinical finding was considered present if a case reported experiencing it during the first 10 days of illness.

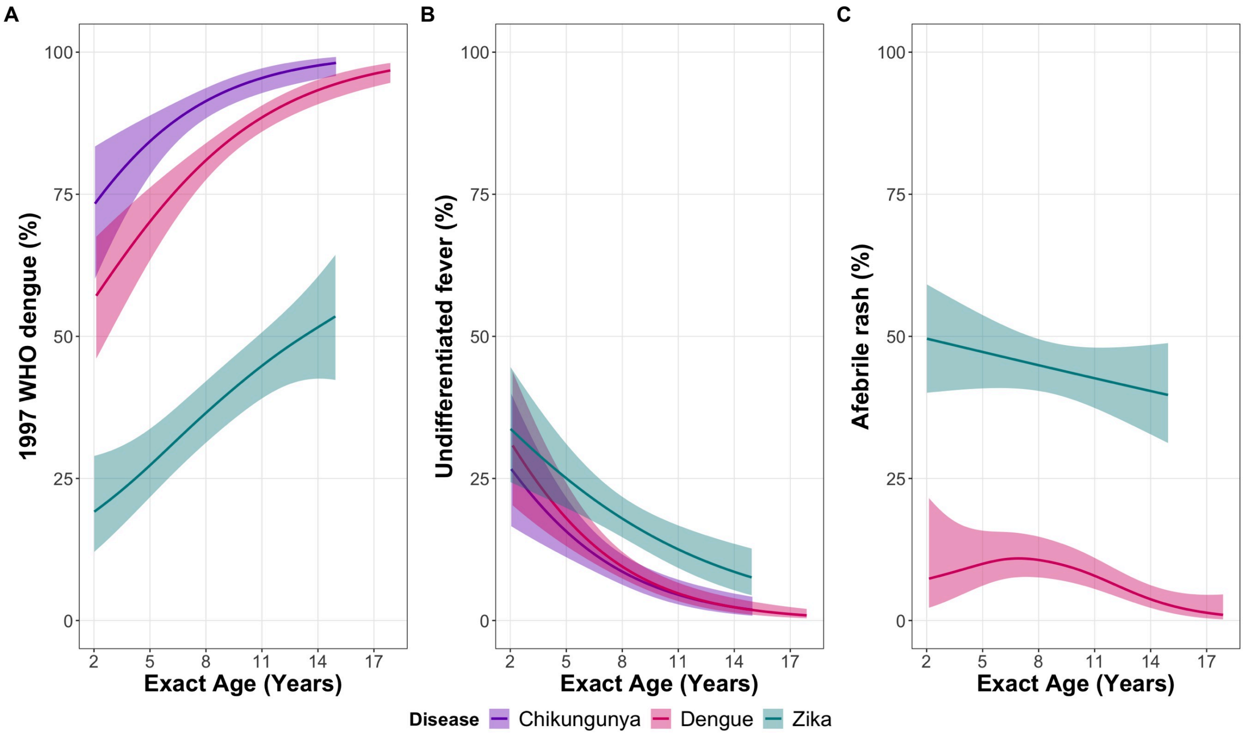

**Figure S3. Age-prevalence trends for dengue, chikungunya, and Zika cases by clinical presentation.** Cases that exhibited clinical findings consistent with one of three broad profiles were tested by diagnostic laboratory methods. The three profiles are: the 1997 WHO case definition for dengue **(A)**; undifferentiated fever without evident cause, with or without any other clinical finding **(B)**; or afebrile rash, with or without any other clinical finding **(C)**. Cases were categorized by these three clinical profiles, and the age-specific prevalence of each clinical profile by disease was estimated using logistic GAMs. Trendlines are superimposed on 95% confidence bands. For example, at 2 years of age, 19% of Zika cases present with a dengue-like clinical profile, 31% with undifferentiated fever, and 50% present with afebrile rash. For this analysis, dengue and Zika cases captured before we started testing suspected cases presenting with afebrile rash were dropped to ensure fair comparisons. All chikungunya cases were captured before the testing criterion for afebrile rash was instituted. However, all chikungunya cases are included for comparison to dengue and Zika cases. Because a recent history of fever or feverishness would suffice to be considered febrile and hence exclude the possibility of a case being classified as experiencing afebrile rash, the use of antipyretic medication would not bias the classification of cases into the three broad profiles.

Abbreviations: GAM, generalized additive model; WHO, World Health Organization

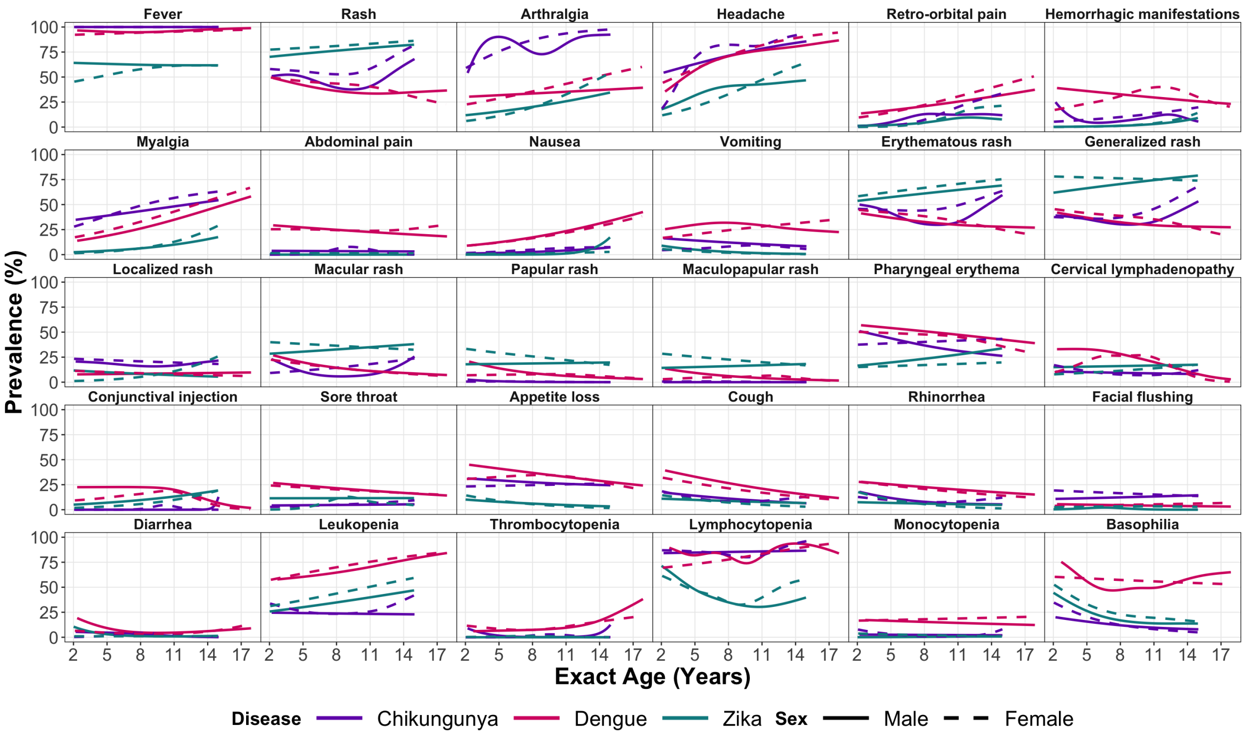

**Figure S4. Age-prevalence trends for clinical features among the dengue, chikungunya, and Zika cases in the PDCS, by sex.** Trend lines were estimated using a logistic GAM. The trend lines may appear less smooth than in Figure 1 because the stratification by sex reduces the sample size that is available to estimate age-prevalence trend lines. 95% confidence bands are not shown to enable a clearer understanding of the underlying trends.

Abbreviations: GAM, generalized additive model; PDCS; Pediatric Dengue Cohort Study

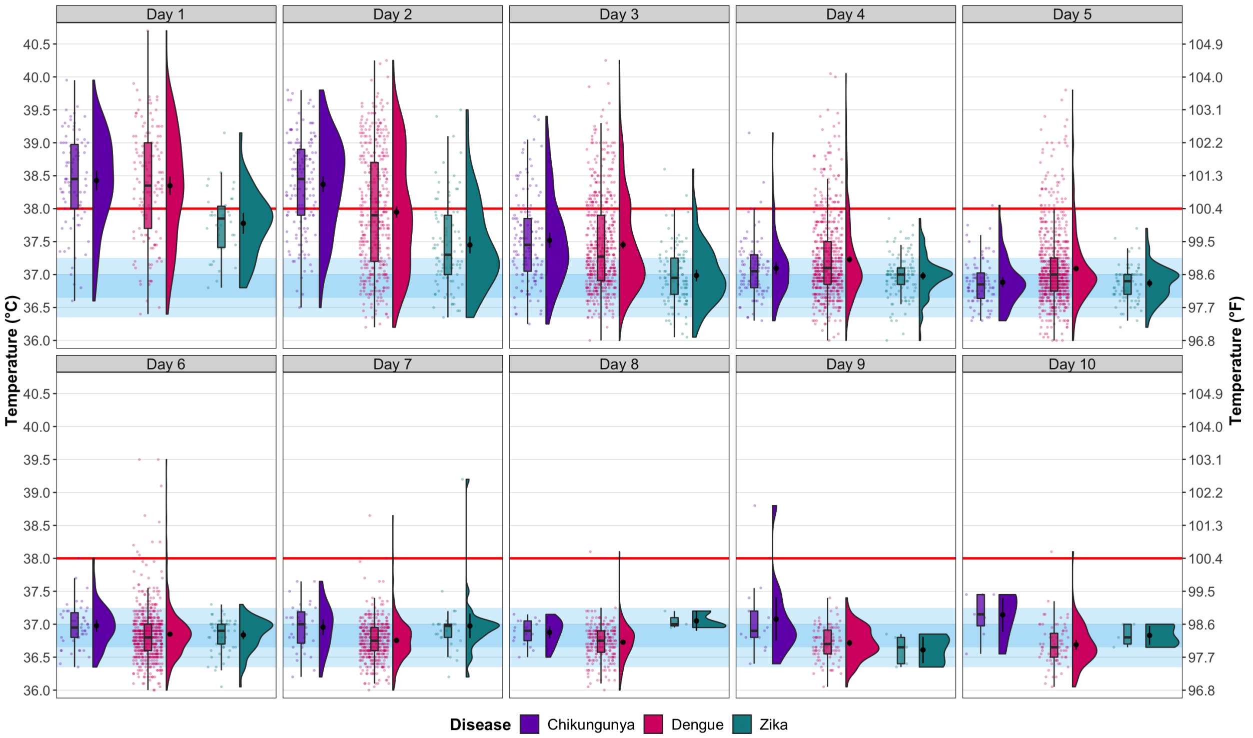

**Figure S5. Raincloud plot of temperature by day and disease among participants not under the influence of antipyretics at the time of the medical consult.** Each PDCS participant presenting to the study health center for evaluation has their temperature taken by a nurse and then by a doctor during the medical consult. Each dot represents the average of these two temperature readings. The data's quartiles and density are summarized by boxplots and violin plots, respectively. Means and 95% CIs, derived from day-and-disease-specific averages, are overlaid on the violin plots. The red line corresponds to our fever threshold of 38·0°C (100·4°F). The blue shaded regions correspond to the 5th, 25th, 75th, and 95th percentile of temperatures (36·35, 36·65, 37·00, 37·25°C; 97·43, 97·97, 98·60, 99·05°F) among healthy PDCS participants taken during 2,341 routine physicals from 2007-2019. This blue shaded region thus represents the distribution of temperature among the PDCS population while healthy. Temperature data was only included for participants who were not on anti-pyretic medication at the time of their medical evaluation.

Abbreviations: CI, confidence interval; PDCS, Pediatric Dengue Cohort Study

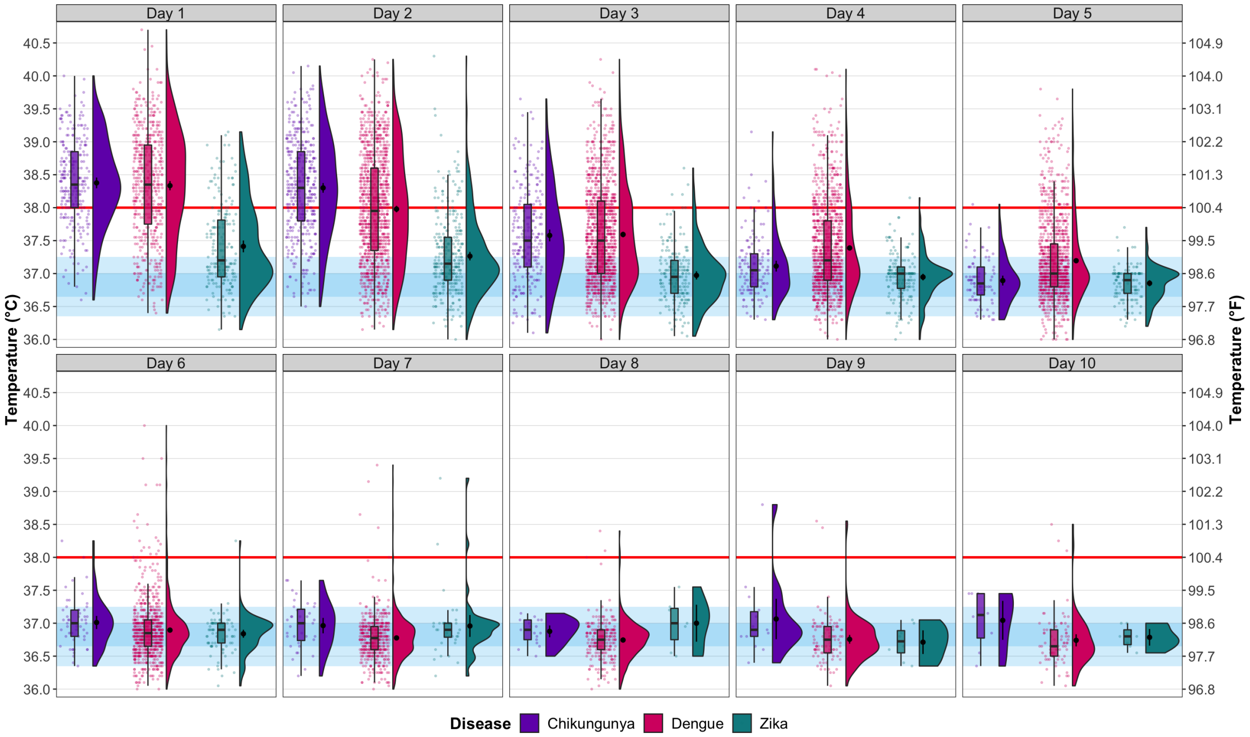

**Figure S6. Raincloud plot of temperature by day and disease among all participants.** Each PDCS participant presenting to the study health center for evaluation has their temperature taken by a nurse and then by a doctor during the medical consult. Each dot represents the average of these two temperature readings. The data's quartiles and density are summarized by boxplots and violin plots, respectively. Means and 95% CIs, derived from day-and-disease-specific averages, are overlaid on the violin plots. The red line corresponds to our fever threshold of 38·0°C (100·4°F). The blue shaded regions correspond to the 5$^{th}$, 25$^{th}$, 75$^{th}$, and 95$^{th}$ percentile of temperatures (36·35, 36·65, 37·00, 37·25°C; 97·43, 97·97, 98·60, 99·05°F) among healthy PDCS participants taken during 2,341 routine physicals from 2007-2019. This blue shaded region thus represents the distribution of temperature among the PDCS population while healthy.

Abbreviations: CI, confidence interval; PDCS, Pediatric Dengue Cohort Study

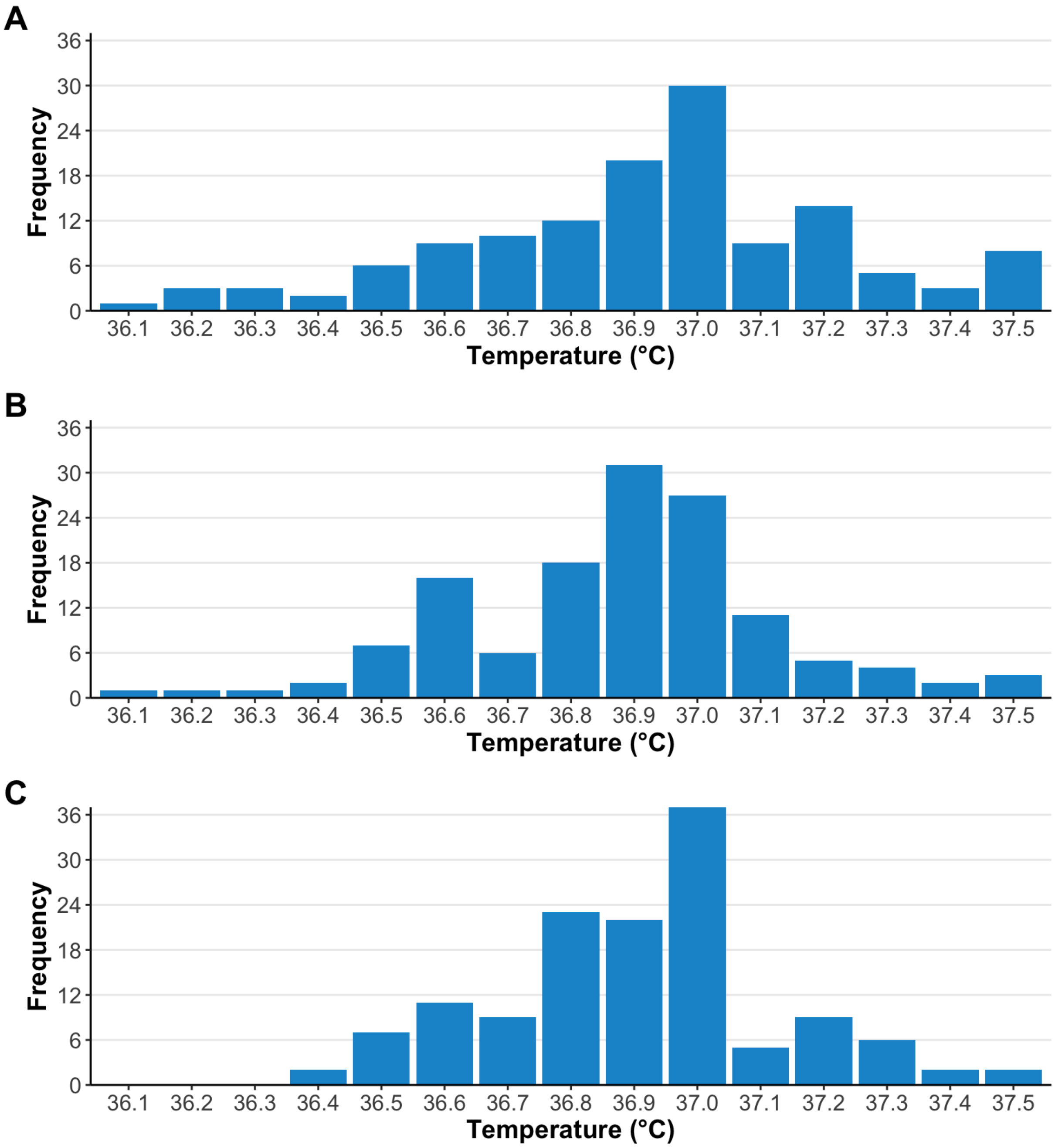

**Figure S7. Temperature readings for the 135 medical consults made by the 62 afebrile dengue cases**. Data shown correspond to temperature readings **(A)** administered by the intake nurse, **(B)** administered by the physician during the medical consult, and **(C)** the average of the two temperature readings. Each figure displays 135 temperature readings. Data corresponding to the per-consult average (C) is used during the analyses given the natural variation across temperature readings; however, the underlying data in (A) and (B) is shown for completion.

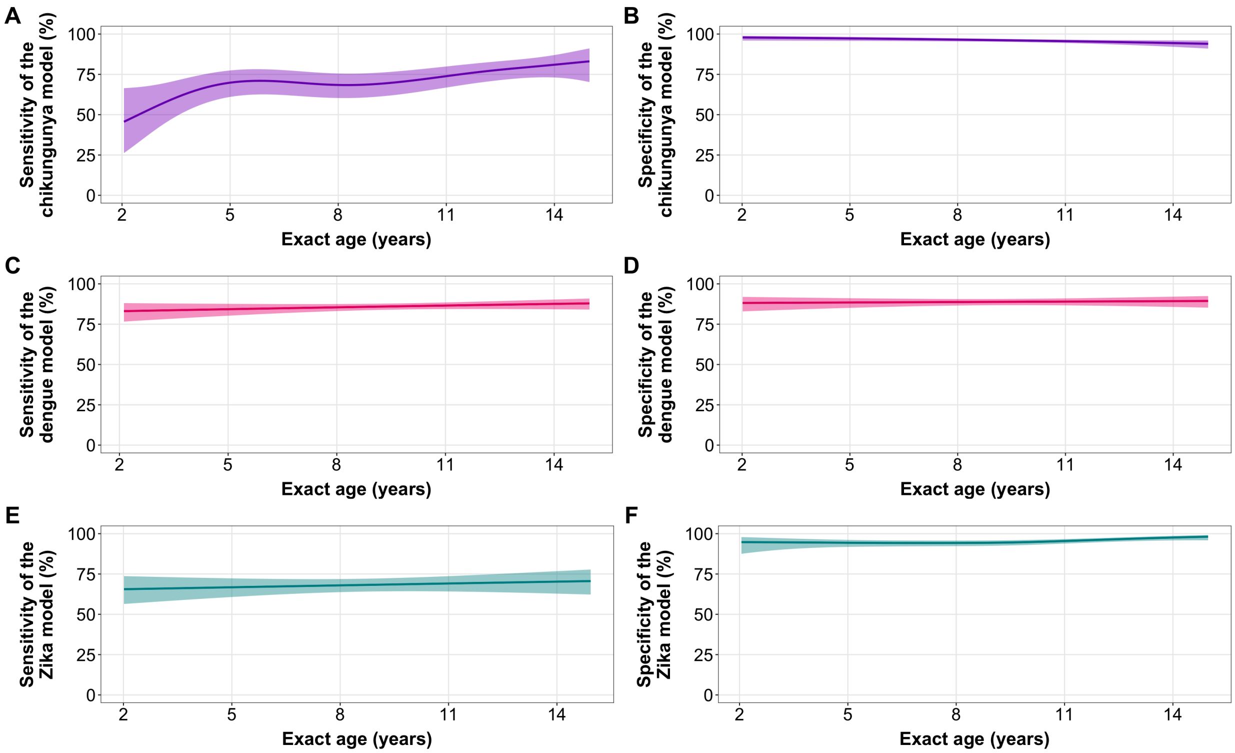

**Figure S8. Sensitivity and specificity plots by age for the chikungunya (A-B), dengue (C-D), and Zika (E-F) full models based on clinical variables from days 1-10 of illness.** Trend lines and 95% confidence bands were estimated with logistic generalized additive models. The models characterized here are the same as in Panel 3A, not models that only include the five most important clinical findings.

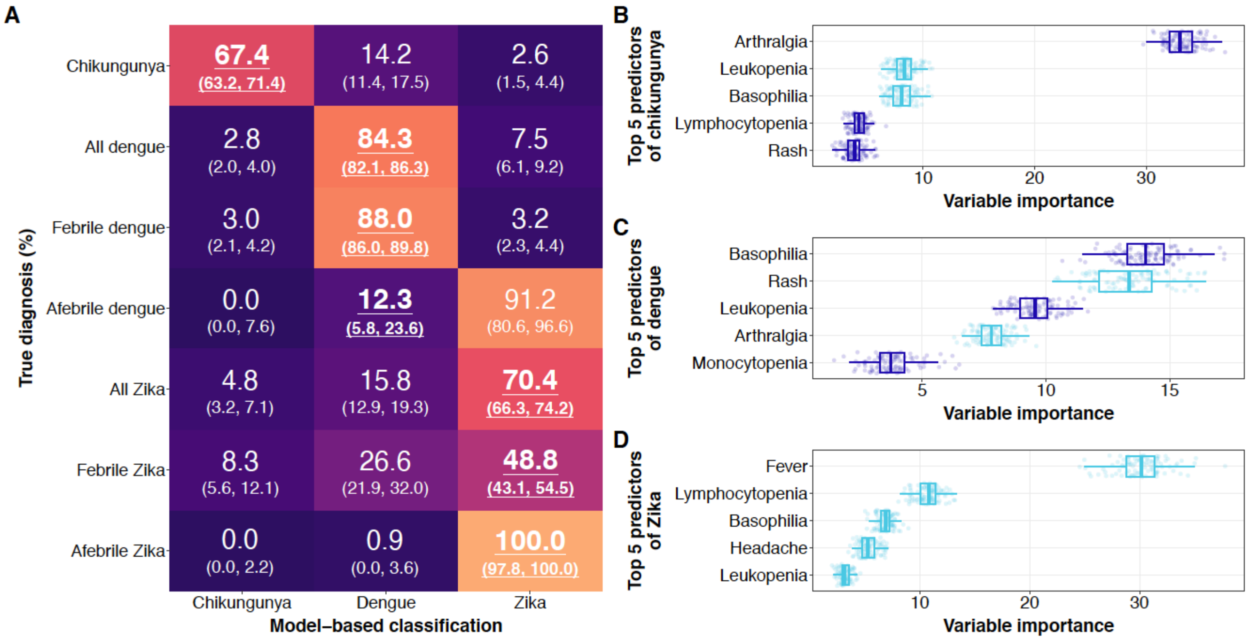

**Figure S9. Reduced machine learning model results for PDCS cases in Managua, Nicaragua (2005-2023) based on all clinical variables across days 1-3 of illness. (A)** Three separate models were constructed to classify, based on the 30 evaluated clinical findings only from days 1-3 of illness, dengue from chikungunya and Zika (the dengue model), chikungunya from dengue and Zika (the chikungunya model), and Zika from dengue and chikungunya (the Zika model). The values shown indicate the percentage of true cases (y-axis) that were classified as a given disease by the disease-specific model (x-axis). For example, 67·4% of chikungunya cases were classified as chikungunya cases by the chikungunya model, and 2·8% of all dengue cases were classified as chikungunya cases by the chikungunya model. Percentages in panel A that are bold and underlined denote correct classifications (which an ideal classifier maximizes), and those in regular font represent incorrect classifications (which an ideal classifier minimizes). Percentages in parentheses denote 95% confidence intervals. Model results are reported for all three diseases and by fever status for dengue and Zika cases. As the testing criteria for all suspected chikungunya cases required fever, we excluded fever from the chikungunya model to limit autocorrelation between predictors and the outcome variable. The chikungunya model had a sensitivity of 67·4% (95% CI: 63·2, 71·4) and an overall specificity of 96·6 (95% CI: 95·6, 97·3). The dengue model had a sensitivity of 84·3% (95% CI: 82·1, 86·3) and an overall specificity of 85·0 (95% CI: 82·6, 87·1). The Zika model had a sensitivity of 70·4% (95% CI: 66·3, 74·2) and an overall specificity of 94·0 (95% CI: 92·7, 95·0). **(B-D)** The five most important clinical findings that helped correctly classify cases are shown for the chikungunya (B), dengue (C), and Zika (D) models. Each dot represents the variable importance value of a given clinical finding across 100 model iterations. Variables further to the right are more important for classification purposes. If the presence of a clinical finding was important to the classification, it is shown in dark blue; if its absence was important, it is shown in light blue. The top five most important variables for each model, paired with information about whether their presence or absence is relevant for classification, together determine how each given variable is helpful in classifying cases. The lower and upper whiskers on the Tukey-style boxplots extend from the first and third quartiles of the data to the smallest and largest values at most 1.5 times the interquartile range, respectively. Data outside these whiskers are classically referred to as outliers.

Abbreviations: PDCS, Pediatric Dengue Cohort Study

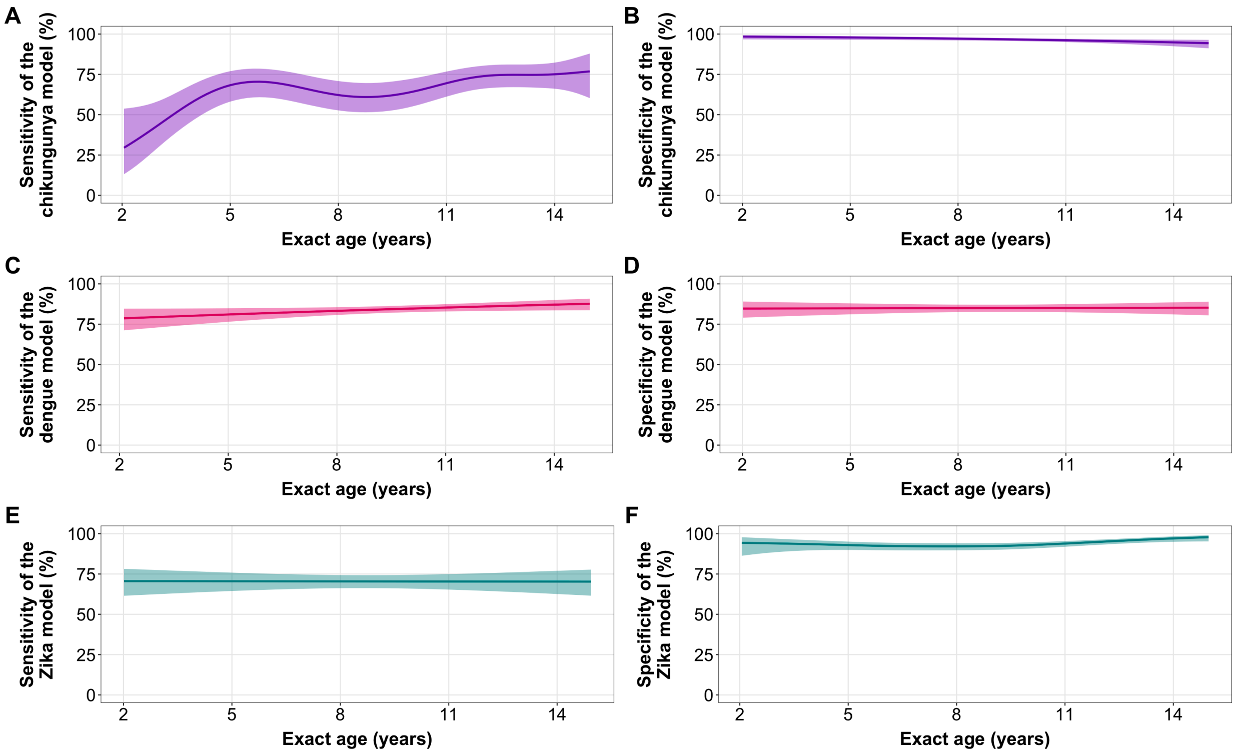

**Figure S10. Sensitivity and specificity plots by age for the chikungunya (A-B), dengue (C-D), and Zika (E-F) reduced models based on clinical variables from days 1-3 of illness.** Trend lines and 95% confidence bands were estimated with logistic generalized additive models. The models characterized here are the same as in Panel S9A, not models that only include the five most important clinical findings.

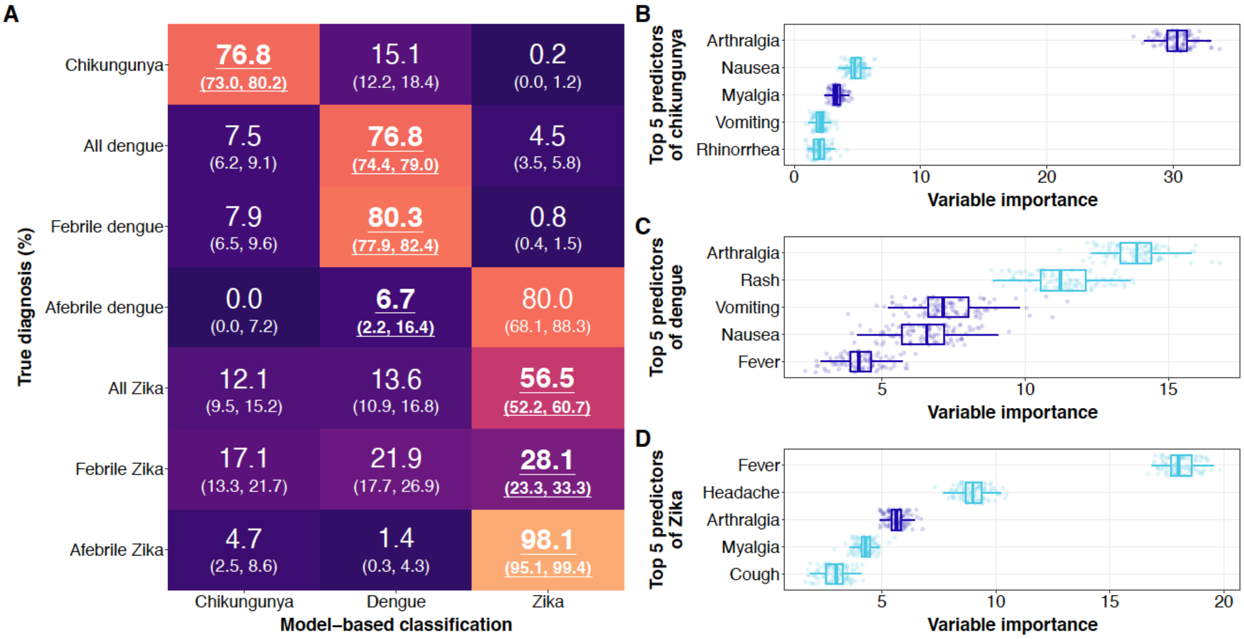

**Figure S11. Reduced machine learning model results for PDCS cases in Managua, Nicaragua (2005-2023) on clinical variables only (no laboratory findings) from days 1-10 of illness. (A)** Three separate models were constructed, based on the 25 clinical findings not derived from complete blood counts, to classify across days 1-10 of illness dengue from chikungunya and Zika (the dengue model), chikungunya from dengue and Zika (the chikungunya model), and Zika from dengue and chikungunya (the Zika model). The values shown indicate the percentage of true cases (y-axis) that were classified as a given disease by the disease-specific model (x-axis). For example, 76·8% of chikungunya cases were classified as chikungunya cases by the chikungunya model, and 7·5% of all dengue cases were classified as chikungunya cases by the chikungunya model. Percentages in panel A that are bold and underlined denote correct classifications (which an ideal classifier maximizes), and those in regular font represent incorrect classifications (which an ideal classifier minimizes). Percentages in parentheses denote 95% confidence intervals. Model results are reported for all three diseases and by fever status for dengue and Zika cases. As the testing criteria for all suspected chikungunya cases required fever, we excluded fever from the chikungunya model to limit autocorrelation between predictors and the outcome variable. The chikungunya model had a sensitivity of 76·8% (95% CI: 73·0, 80·2) and an overall specificity of 91·2 (95% CI: 89·8, 92·4). The dengue model had a sensitivity of 76·8% (95% CI: 74·4, 79·0) and an overall specificity of 85·7 (95% CI: 83·4, 87·7). The Zika model had a sensitivity of 56·5% (95% CI: 52·2, 60·7) and an overall specificity of 96·7 (95% CI: 95·8, 97·4). **(B-D)** The five most important clinical findings that helped correctly classify cases are shown for the chikungunya (B), dengue (C), and Zika (D) models. Each dot represents the variable importance value of a given clinical finding across 100 model iterations. Variables further to the right are more important for classification purposes. If the presence of a clinical finding was important to the classification, it is shown in dark blue; if its absence was important, it is shown in light blue. The top five most important variables for each model, paired with information about whether their presence or absence is relevant for classification, together determine how each given variable is helpful in classifying cases. The lower and upper whiskers on the Tukey-style boxplots extend from the first and third quartiles of the data to the smallest and largest values at most 1.5 times the interquartile range, respectively. Data outside these whiskers are classically referred to as outliers.

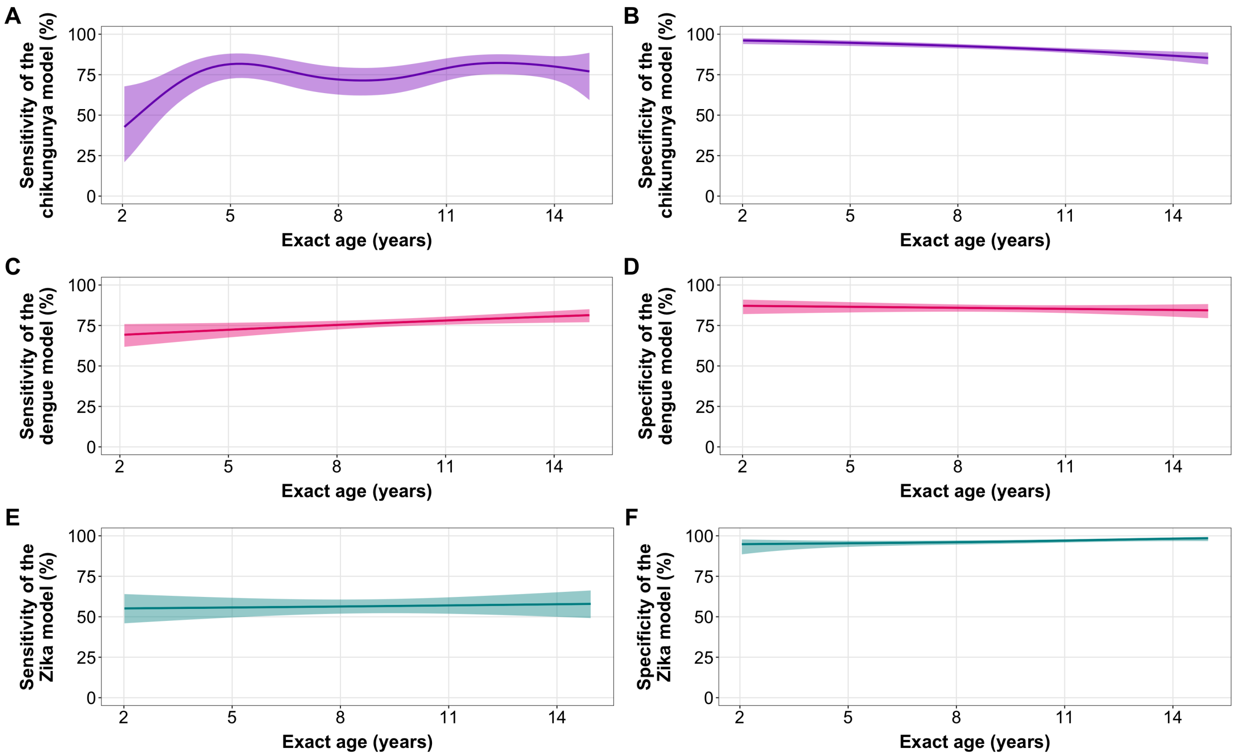

**Figure S12. Sensitivity and specificity plots by age for the chikungunya (A-B), dengue (C-D), and Zika (E-F) reduced models based on clinical variables only (no laboratory findings) across days 1-10 of illness.** Trend lines and 95% confidence bands were estimated with logistic generalized additive models. The models characterized here are the same as in Panel S11A, not models that only include the five most important clinical findings.

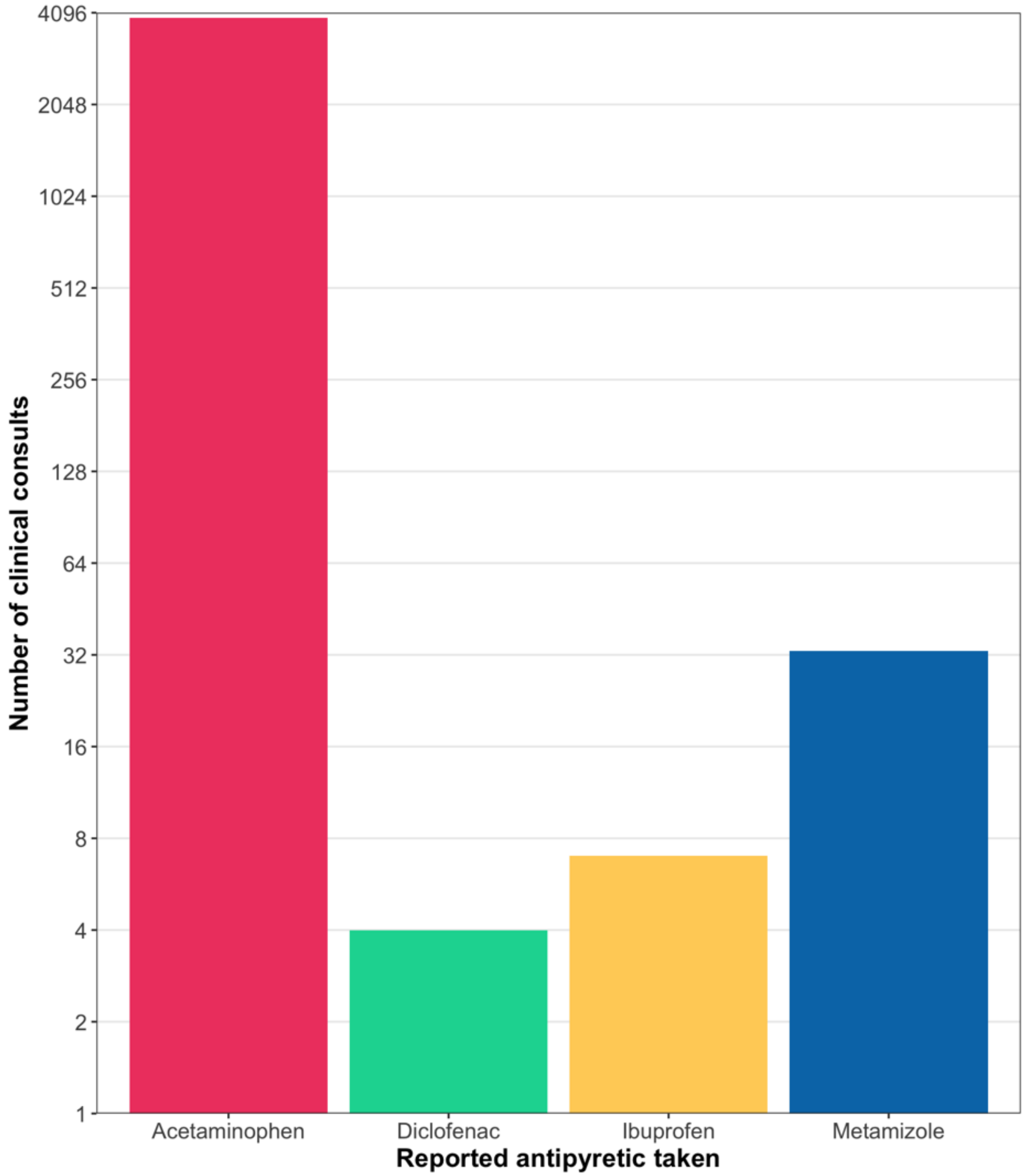

**Figure S13. Antipyretic medication reported by PDCS patients and caregivers at the time of the medical consult.** The y-axis is on a log 2 scale.

Abbreviations: PDCS, Pediatric Dengue Cohort Study

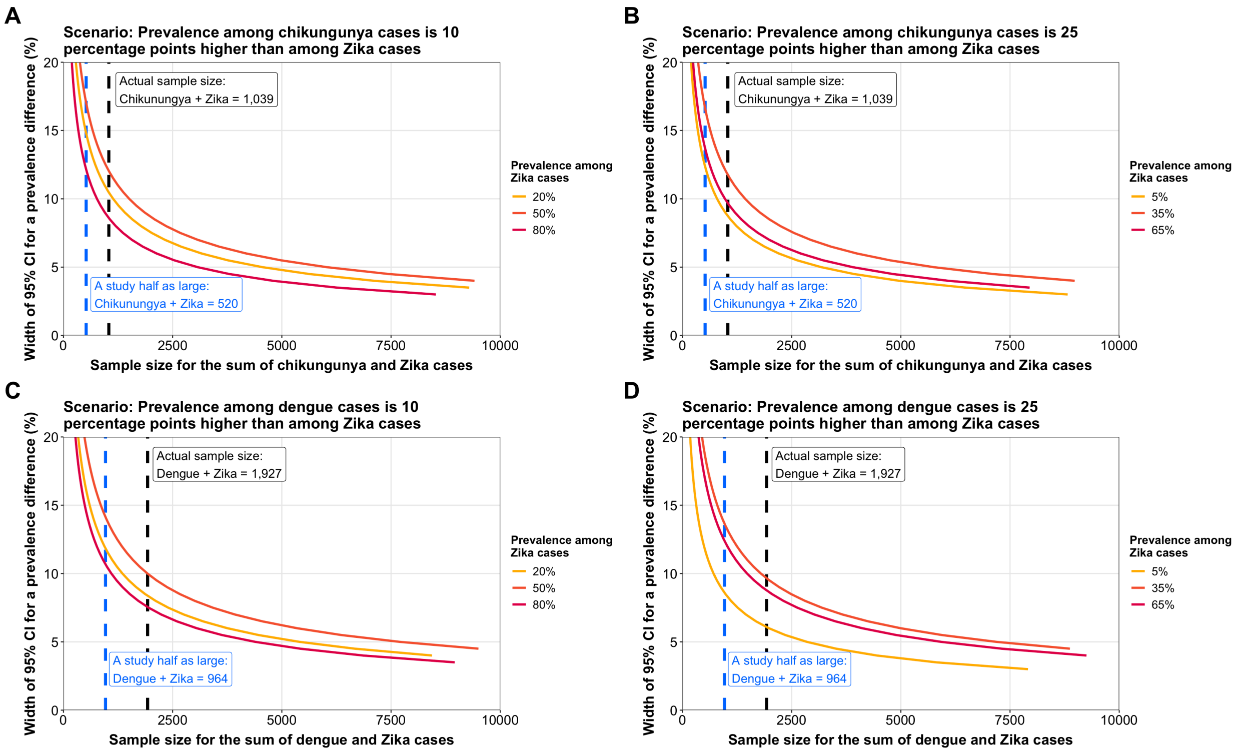

**Figure S14. Precision-based, post-hoc sample size calculations based on the width of a 95% confidence interval for the prevalence difference.** Comparisons are shown for chikungunya vs. Zika (A-B) and dengue vs. Zika cases (C-D). Since the number of Zika cases (N=522) is approximately that of the number of chikungunya cases (N=517), analyses of dengue vs. chikungunya would be duplicative with the latter scenario. Precision-based sample size calculations vary based on the difference between the prevalence of a given disease among two types of cases (this difference is the effect size in traditional power calculations) and the ratio of the two types of cases. Therefore, we estimated precision-based sample size calculations to account for all these factors. **(A)** Estimates assuming the prevalence of a given disease is 10 percentage points higher among chikungunya than Zika cases. The three scenarios in panel A represents a prevalence difference of 10% where the underlying prevalences are low, medium, or high. For example, the yellow line assumes a prevalence of 30% among chikungunya cases and a prevalence of 20% among Zika cases. The orange line assumes a prevalence of 60% among chikungunya cases and 50% among Zika cases. The red line assumes a prevalence of 90% among chikungunya cases and 80% among Zika cases. **(B)** Estimates assuming the prevalence of a given disease is 25 percentage points higher among chikungunya than Zika cases. **(C)** Estimates assuming the prevalence of a given disease is 10 percentage points higher among dengue than Zika cases. **(D)** Estimates assuming the prevalence of a given disease is 25 percentage points higher among dengue than Zika cases. For all scenarios, 1) the ratio of the number of cases represents the actual, observed ratio (*e.g.*, in the comparisons of chikungunya to Zika cases in panels A-B, the ratio of Zika to chikungunya cases was set to 522/517, reflecting their actual sample size); 2) three estimates of the precision-sample size association are given assuming the underlying prevalence of a given disease among the cases is low (yellow), medium (orange), or high (red); 3) sample size estimates were generated from a precision (interval width) of 1 to 20 percentage points in increments of 0.5 points; 4) the maximum sample size of the participants in the comparison of interest is set to 10,000, such that curves are truncated at the nearest estimated result prior to the overall sample size of interest exceeding 10,000; and 5) the black dashed line represents the actual sample size and precision achieved in the present study, whereas the blue dashed line represents the sample size and precision that would be obtained from a hypothetical study half the size of the present study.

### SUPPLEMENTAL REFERENCES

1. Kuan G, Ramirez S, Gresh L, Ojeda S, Melendez M, Sanchez N, et al. Seroprevalence of anti-chikungunya virus antibodies in children and adults in Managua, Nicaragua, after the first chikungunya epidemic, 2014-2015. Bingham A, editor. PLoS Negl Trop Dis. 2016 Jun 20;10(6):e0004773.
2. Kuan G, Gordon A, Aviles W, Ortega O, Hammond SN, Elizondo D, et al. The Nicaraguan Pediatric Dengue Cohort Study: Study design, methods, use of information technology, and extension to other infectious diseases. Am J Epidemiol. 2009 Jul 1;170(1):120–9.
3. Balmaseda A, Standish K, Mercado JCC, Matute JCC, Tellez Y, Saborío S, et al. Trends in patterns of dengue transmission over 4 years in a pediatric cohort study in Nicaragua. J Infect Dis. 2010 Jan 1;201(1):5–14.
4. World Health Organization. Dengue: Guidelines for Diagnosis, Treatment, Prevention and Control (New Edition 2009). World Health Organization; 2009. 158 p.
5. Waggoner JJ, Gresh L, Mohamed-Hadley A, Ballesteros G, Davila MJV, Tellez Y, et al. Single-reaction multiplex reverse transcription PCR for detection of Zika, chikungunya, and dengue viruses. Emerg Infect Dis. 2016 Jul;22(7):1295–7.
6. Santiago GA, Vázquez J, Courtney S, Matías KY, Andersen LE, Colón C, et al. Performance of the Trioplex real-time RT-PCR assay for detection of Zika, dengue, and chikungunya viruses. Nat Commun. 2018 Dec 11;9(1):1391.
7. Lanciotti RS, Kosoy OL, Laven JJ, Velez JO, Lambert AJ, Johnson AJ, et al. Genetic and serologic properties of Zika virus associated with an epidemic, Yap State, Micronesia, 2007. Emerg Infect Dis. 2008 Aug;14(8):1232–9.
8. Waggoner JJ, Ballesteros G, Gresh L, Mohamed-Hadley A, Tellez Y, Sahoo MK, et al. Clinical evaluation of a single-reaction real-time RT-PCR for pan-dengue and chikungunya virus detection. J Clin Virol. 2016 May;78:57–61.
9. Burger-Calderon R, Bustos Carrillo F, Gresh L, Ojeda S, Sanchez N, Plazaola M, et al. Age-dependent manifestations and case definitions of paediatric Zika: a prospective cohort study. Lancet Infect Dis. 2020 Dec;20(3):371–80.
10. Gordon A, Gresh L, Ojeda S, Katzelnick LC, Sanchez N, Mercado JC, et al. Prior dengue virus infection and risk of Zika: A pediatric cohort in Nicaragua. von Seidlein L, editor. PLOS Med. 2019 Jan 22;16(1):e1002726.
11. Katzelnick LC, Gresh L, Halloran ME, Mercado JC, Kuan G, Gordon A, et al. Antibody-dependent enhancement of severe dengue disease in humans. Science. 2017 Nov 17;358(6365):929–32.
12. World Health Organization. Dengue haemorrhagic fever: Diagnosis, treatment, prevention, and control. 2nd ed. Geneva; 1997.
13. Pan American Health Organization. Case definitions, clinical classification, and disease phases: Dengue, Chikungunya, and Zika. Washington, DC; 2023.
14. World Health Organization - Regional Office for South-East Asia. Guidelines for prevention and control of Chikungunya fever. 2009. p. 19.
15. PAHO/WHO. Zika resources: Case definitions. 2016.
16. World Health Organization. Zika virus disease - Interim case definition. World Health Organization; 2016.
17. Balmaseda A, Hammond SN, Pérez L, Tellez Y, Saborío SI, Mercado JC, et al. Serotype-specific differences in clinical manifestations of dengue. Am J Trop Med Hyg. 2006 Mar;74(3):449–56.
18. Narvaez F, Montenegro C, Juarez JG, Zambrana JVJV, Gonzalez K, Videa E, et al. Dengue severity by serotype and immune status in 19 years of pediatric clinical studies in Nicaragua. Marques ETA, editor. PLoS Negl Trop Dis. 2025 Jan 10;19(1):e0012811.
19. Hastie T, Tibshirani R. Generalized Additive Models. Stat Sci. 1986;1(3):297–318.
20. Wood SN. Generalized Additive Models: An Introduction with R. 2nd ed. Chapman and Hall / CRC Press; 2017.
21. Wood SN. Fast stable restricted maximum likelihood and marginal likelihood estimation of semiparametric generalized linear models. J R Stat Soc Ser B (Statistical Methodol. 2011 Jan;73(1):3–36.
22. Wood SN. Stable and Efficient Multiple Smoothing Parameter Estimation for Generalized Additive Models. J Am Stat Assoc. 2004 Sep;99(467):673–86.
23. Wood SN, Fasiolo M. A generalized Fellner-Schall method for smoothing parameter optimization with application to Tweedie location, scale and shape models. Biometrics. 2017 Dec 1;73(4):1071–81.

24. Campbell I. Chi-squared and Fisher–Irwin tests of two-by-two tables with small sample recommendations. Stat Med. 2007 Aug 30;26(19):3661–75.
25. Miettinen O, Nurminen M. Comparative analysis of two rates. Stat Med. 1985;4(2):213–26.
26. Stevenson M, Nunes T, Heuer C, Marshall J, Sanchez J, Thorton R, et al. epiR: Tools for the Analysis of Epidemiological Data. 2018.
27. PAHO. Guidelines for the clinical diagnosis and treatment of dengue, chikungunya, and Zika. Washington, D.C.: Pan American Health Organization; 2022.
28. Elith J, Leathwick JR, Hastie T. A working guide to boosted regression trees. J Anim Ecol. 2008 Jul 1;77(4):802–13.
29. Ridgeway G, Developers G. gbm: Generalized Boosted Regression Models. 2024.
30. Becker DJ, Albery GF, Sjodin AR, Poisot T, Bergner LM, Chen B, et al. Optimising predictive models to prioritise viral discovery in zoonotic reservoirs. The Lancet Microbe. 2022 Aug 1;3(8):e625–37.
31. Mull N, Carlson CJ, Forbes KM, Becker DJ. Virus isolation data improve host predictions for New World rodent orthohantaviruses. J Anim Ecol. 2022 Jun 1;91(6):1290–302.
32. Gholamy A, Kreinovich V, Kosheleva O. Why 70/30 or 80/20 Relation Between Training and Testing Sets: A Pedagogical Explanation. Dep Tech Reports. 2018 Feb 1.
33. Freeman E, Moisen G. PresenceAbsence: An R package for presence-absence model analysis. J Stat Softw. 2008;23(11):1–31.
34. Barrett M. precisely: Estimate sample size based on precision rather than power. 2021.
35. Rothman KJ, Greenland S. Planning study size based on precision rather than power. Epidemiology. 2018;29(5):599–603.
36. Wickham H. ggplot2: Elegant Graphics for Data Analysis. Springer-Verlag New York; 2016.
37. Wickham H. ggplot2. 1st ed. New York City: Springer; 2009. 1–212 p.
38. Pedersen TL. patchwork: The Composer of Plots. 2019.
39. RStudio Team. RStudio: Integrated Development for R. Boston: RStudio, Inc.; 2016.
40. R Core Team. R: A language and environment for statistical computing. R Foundation for Statistical Computing. Vienna: R Foundation for Statistical Computing; 2019.
41. Wickham H, Bryan J. readxl: Read Excel Files. 2018.
42. Schwartz M. WriteXLS: Cross-Platform Perl Based R Function to Create Excel 2003 (XLS) and Excel 2007 (XLSX) Files. 2015.
43. Wickham H, François R, Henry L, Müller K. dplyr: A Grammar of Data Manipulation. 2018.
44. Harrell F, Dupont C. Hmisc: Harrell Miscellaneous. 2020.
45. Mahmoudain M. varhandle: Functions for Robust Variable Handling. 2020.
46. Bache SM, Wickham H. magrittr: A Forward-Pipe Operator for R. 2014.
